# Associations between corticolimbic glutamatergic metabolites and functional connectivity in people at clinical high-risk for psychosis

**DOI:** 10.64898/2026.04.08.26350385

**Authors:** Abigail Gee, Nicholas R. Livingston, Amanda Kiemes, Samuel R. Knight, Paulina B. Lukow, David J. Lythgoe, Natasha Vorontsova, Jacek Donocik, James Davies, Eugenii A. Rabiner, Federico Turkheimer, Matthew B. Wall, Thomas J. Spencer, Andrea de Micheli, Paolo Fusar-Poli, Anthony A. Grace, Steven C. Williams, Philip McGuire, Paola Dazzan, Gemma Modinos

**Affiliations:** Department of Psychological Medicine, Institute of Psychiatry, Psychology & Neuroscience, King’s College London, London, United Kingdom; Artificial Intelligence Centre for Value-based Healthcare, London, United Kingdom; Institute of Cognitive Neuroscience, University College London, London, United Kingdom; Department of Neuroimaging, Institute of Psychiatry, Psychology & Neuroscience, King’s College London, London, United Kingdom; Southwark Early Intervention Service, South London and the Maudsley NHS Foundation Trust, London, United Kingdom; Lewisham Early Intervention Service, South London and the Maudsley NHS Foundation Trust, London, United Kingdom; Perceptive Inc., Burlington Danes Building, Imperial College London, London, United Kingdom; Department of Psychosis Studies, Institute of Psychiatry, Psychology & Neuroscience, King’s College London, London, United Kingdom; Outreach and Support in South London (OASIS) Service, South London and the Maudsley NHS Foundation Trust, London, United Kingdom; Department of Brain and Behavioural Sciences, University of Pavia, Pavia, Italy; Departments of Neuroscience, Psychiatry and Psychology, University of Pittsburgh, Pittsburgh, United States; Department of Psychiatry, University of Oxford, Oxford, United Kingdom; NIHR Oxford Health Biomedical Research Centre, University of Oxford, Oxford, United Kingdom; MRC Centre for Neurodevelopmental Disorders, Institute of Psychiatry, Psychology & Neuroscience, King’s College London, London, United Kingdom; NIHR Maudsley Biomedical Research Centre, King’s College London, London, United Kingdom

**Author notes:** **Corresponding author:** Prof. Gemma Modinos, Department of Psychological Medicine, Institute of Psychiatry, Psychology & Neuroscience, 16 De Crespigny Park, King’s College London, London, SE5 8AF, UK.

## Abstract

Recent evidence suggests that psychosis involves glutamatergic dysfunction and altered activity/connectivity within corticolimbic circuitry. While altered relationships between corticolimbic glutamatergic metabolite levels and resting-state functional connectivity (FC) have been described in schizophrenia and first-episode psychosis (FEP), whether these disruptions are also present prior to psychosis onset remains unclear. We measured Glx (glutamate + glutamine) levels in the anterior cingulate cortex (ACC) and hippocampus with magnetic resonance spectroscopy (MRS), and resting-state FC between corticolimbic regions of interest (ACC, hippocampus, amygdala and nucleus accumbens (NAc)) in antipsychotic-naïve participants at clinical high-risk for psychosis (CHR-P, n=22), compared to healthy controls (HC, n=23) and FEP participants (n=10). Primary analyses compared corticolimbic Glx-FC interactions between CHR-P and HC groups. FEP individuals were included in secondary Glx comparisons but were excluded from FC analyses due to insufficient sample size after quality control. There was a significant interaction between group and ACC Glx for FC between the NAc and the bilateral amygdala and hippocampus (p-FDR=0.021), which was driven by a significant negative association in the CHR-P group (p-FDR=0.005). Complementary seed-to-whole-brain analyses revealed additional negative associations between ACC Glx and FC with the left middle temporal gyrus, and between hippocampal Glx and FC with the parahippocampal and temporal fusiform cortices in CHR-P individuals, which were absent in HC. FEP showed higher Glx than HC across both regions (p=0.015), but there were no significant Glx differences between CHR-P and HC. These data suggest that increased risk for psychosis is associated with altered relationships between corticolimbic connectivity and glutamatergic function.

## Introduction

Glutamatergic dysfunction has been consistently implicated in the pathophysiology of psychosis [1]. In individuals with schizophrenia, this has predominantly involved measuring regional levels of glutamate and glutamine, or their combined measure Glx, non-invasively in vivo using proton magnetic spectroscopy (^1^H-MRS). However, the directionality and location of findings have not been entirely consistent [2], and both antipsychotic treatment [3,4], and degree of treatment response [5–8] can influence glutamatergic metabolite measurements. Focusing on the at-risk and early psychosis stages therefore provides a unique opportunity to characterise glutamatergic function with fewer confounding influences from medication and illness chronicity. Furthermore, single-voxel ^1^H-MRS studies cannot capture the proposed distributed and interconnected nature of these proposed glutamatergic alterations in psychosis. Multimodal studies incorporating multiple voxel locations and exploring the associations with functional connectivity may be a more suitable approach to understand how regional neurochemical alterations relate to wider network dysfunction in early psychosis.

Several meta-analyses of ^1^H-MRS studies have reported differences in glutamatergic metabolite levels in schizophrenia, first episode psychosis (FEP) and individuals at clinical high-risk for psychosis (CHR-P) [2,9–14]. For instance, in schizophrenia there have been reports of higher glutamate and Glx levels in the basal ganglia, higher glutamine in the thalamus, higher Glx in the medial temporal lobe [11], and lower glutamate in the medial frontal cortex [15]. In FEP and CHR-P ^1^H-MRS studies, meta-analytic evidence has highlighted elevations in medial frontal Glx in CHR-P, but no significant differences in regional Glx, glutamate or glutamine in FEP compared to controls [11]. A more recent meta-analysis focusing on those at-risk for psychosis reported higher thalamic Glx concentrations in individuals at genetic risk, but no differences for CHR-P compared to controls in the prefrontal cortex or hippocampus [10]. As the brain’s principal excitatory neurotransmitter, glutamate plays a central role in regulating FC by driving the activation of pyramidal neurons, which form long-range connections between brain regions [16]. Although group differences in regional glutamatergic metabolites are inconsistent, exploring the association with functional connectivity (FC) may reveal mechanistic links underlying symptom development.

Functional magnetic resonance imaging (fMRI) studies have found widespread changes in FC within and between large-scale brain networks such as the default mode (DMN), central executive and salience networks in schizophrenia [17] and FEP samples [18]. There are similar, though less pronounced, alterations reported in CHR-P studies [19]. Corticolimbic structures such as the hippocampus and ACC act as highly connected hubs within these networks, integrating sensory, emotional and cognitive information [20]. Their dense connectivity with other regions, including the amygdala and nucleus accumbens (NAc), places them at a critical interface where glutamatergic disruption can propagate to wider circuits involved in salience processing, reward learning and emotion-cognition integration [21–23]. Hence, glutamatergic dysregulation within corticolimbic hubs may drive circuit and broader network-level connectivity changes, potentially contributing to the emergence of clinical symptoms.

The interaction between regional glutamate and network function is further supported by multimodal neuroimaging. In healthy individuals, a meta-analysis of multimodal ^1^H-MRS-fMRI studies reported that higher regional glutamate concentrations are associated with greater activation in distal, but not local, brain regions [24]. A recent systematic review found that, across six studies, associations between ACC glutamatergic metabolites and resting-state FC are strong and regionally specific in healthy individuals, but consistently weaker or absent in schizophrenia and FEP [25]. Two hippocampal ^1^H-MRS studies included in the same review reported that, in HCs, higher hippocampal glutamate was positively associated with increased connectivity to DMN regions, while in FEP and schizophrenia the strength and direction of these associations were weaker or reversed [25]. To our knowledge, no studies have examined the relationship between glutamatergic metabolites and resting-state FC in CHR-P individuals. Examining the relationship between neurochemical and functional measures prior to the influence of antipsychotic exposure or illness progression could help to disentangle the complex underlying neurobiological mechanisms and to identify potential markers of circuit vulnerability in psychosis risk.

Therefore, this multi-modal neuroimaging study aimed to examine whether the CHR-P state is associated with interactions between Glx concentrations in ACC and hippocampus and resting-state FC in key corticolimbic regions (ACC, hippocampus, amygdala, nucleus accumbens) [20,22]. We hypothesised that the relationship between these measures would be altered in CHR-P compared to HC. Secondary analyses examined group differences in the individual imaging modalities separately (MRS and resting-state fMRI). Finally, exploratory analyses examined whether Glx levels and Glx-FC associations would be related to positive, negative and cognitive symptoms.

## Methods

### Study procedures and participants

The present study used unpublished resting-state fMRI and MRS data from 24 CHR-P individuals, 10 people with first episode psychosis (FEP) and 24 healthy controls (HC) from a wider PET-MR study. Full details of recruitment criteria for the wider PET-MR study have been reported previously [26]. Briefly, the CHR-P and FEP individuals were recruited through South London and the Maudsley (SLaM) Foundation Trust clinical teams, including the early-intervention for psychosis teams and Outreach and Support in South London (OASIS) service [27]. The HC participants were recruited through university circulars and public advertisement from the South London area.

All participants had capacity to consent, were 18-40 years old and had an IQ ≥70 as estimated by an abbreviated version of the Wechsler Adult Intelligence Scale (WAIS-III) [28,29]. Participants had no history of neurological conditions, past or current substance abuse, no MRI contraindications and were not pregnant or breastfeeding. Participants were excluded if they were currently exposed to any medications with prominent GABAergic or glutamatergic mechanisms of action, but we included those taking antidepressant medications. Additional inclusion criteria for HC participants were no self-reported Axis-I mental health disorder or first-degree family history of any mental health disorder.

CHR-P participants had current attenuated symptoms as defined by the Comprehensive Assessment for At-Risk Mental States (CAARMS) [30] and all were antipsychotic naïve. FEP participants had an ICD-10 diagnosis of a psychotic disorder (F20-F29 and F31) [31], rated at least one moderate severity score on the positive subscale of the Positive and Negative Syndrome Scale (PANSS) [32] and were within 2 years of first diagnosis of psychosis. FEP participants were included if they were either antipsychotic naïve, were not currently taking antipsychotics, or were on a stable dose of any antipsychotic (other than clozapine). We aimed to recruit 20 FEP individuals, but due to the impact of the COVID-19 pandemic on early-intervention services and recruitment for research studies, imaging data were ultimately only acquired in 10 FEP. The study was approved by the London/Surrey Research Ethics Committee (17/LO/1130). All participants provided written informed consent before participation in accordance with the Declaration of Helsinki.

### Demographic and clinical data

We collected information on psychosis positive and negative symptom severity (CAARMS in CHR-P and HC, PANSS in FEP), anxiety and depression symptom severity (Hamilton Anxiety (HAM-A) and Depression (HAM-D) Rating Scales), current overall functioning (World Health Organization Disability Assessment Schedule (WHODAS-II) [33] and Social and Occupational Functioning Assessment Scale (SOFAS) [34]), and current psychotropic medications.

### MRI acquisition

This study analysed resting-state fMRI and ^1^H-MRS data acquired during the same scanning session, using a Signa^TM^ simultaneous PET-MR General Electric (GE) scanner with a 12-channel head coil at the Perceptive (formerly, Invicro) Imaging Centre, London, UK. Details of scan acquisition parameters are described in the supplementary materials (supplementary methods 1.1).

### Neuroimaging data processing

#### ^1^H-MRS preprocessing and quality control

^1^H-MRS data was pre-processed and analysed using Osprey version 2.9.0 (Matlab R2024b) following the standard procedures for data processing, quantification, and modelling using Osprey’s built in linear combination algorithm [35]. A vendor and sequence matched metabolite basis set was used for quantification (supplementary methods 1.2).

Quality control of the spectra involved assessment of the signal-to-noise ratio (SNR), spectral linewidth (full width half-maximum, FWHM), and visual inspection. Spectra with SNR<2 standard deviations above the mean or FWHM>2 standard deviations above the mean were removed from subsequent analysis [36]. We assessed and reported relative fit quality calculated as the ratio of the fit residual amplitude to the standard deviation of the noise. The ACC ^1^H-MRS data were missing for two HC and one CHR-P participant, and data were excluded due to insufficient spectral quality for two HC and one CHR-P participant. The hippocampal ^1^H-MRS data were missing for one HC and three CHR-P participants and were excluded due to insufficient quality for one HC and two CHR-P participants. For ^1^H-MRS data analyses we therefore included 19 HC, 18 CHR-P and 10 FEP participants with usable hippocampal and ACC data (supplementary table 1 (ST1)).

Structural T1 images were used to spatially map the voxel location onto each participant’s native anatomical space and to extract voxel-specific tissue fractions for gray matter, white matter and cerebral spinal fluid. Co-registration was performed using Osprey’s built-in co-registration module and segmentation was performed using SPM12. Voxel placement was verified by visual inspection of the voxel mask in subject space and plotting of normalised voxel locations.

Water-scaled tissue-corrected Glx values in molar units (mmol/kg) were used for subsequent analyses. Further details on ^1^H-MRS methodology are provided in the supplementary methods (section 1.2), including the MRSinMRS checklist.

#### Resting-state fMRI preprocessing and quality control

The resting-state fMRI (rs-fMRI) data was preprocessed using fMRIPrep 25.0.0 [37] and CONN 22.v2507 [38]. Structural T1 images were bias field corrected, skull-stripped, segmented and normalised to MNI space. Functional data were co-registered, spatially smoothed (6mm FWHM), and denoised using CONN’s standard pipeline (including aCompCor, motion regression, despiking, scrubbing and band-pass filtering of 0.008–0.09 Hz). Participants with mean framewise displacement (FD) > 0.5mm were excluded. Full preprocessing details are provided in the supplementary methods (section 1.3).

All participants had rs-fMRI data but two CHR-P participants and three FEP participants were excluded due to head motion. The FEP group was excluded from the rs-fMRI-MRS analyses to n=7 after quality control, which was considered insufficiently powered for analyses combining imaging modalities. Analyses therefore included 20 HC and 20 CHR-P participants for the ACC dataset, and 22 HC and 18 CHR-P participants for the hippocampal dataset (ST1).

#### Generation of seed and region-of-interest (ROI) masks

Subject-specific left hippocampus and ACC seed masks were defined using the normalised MRS masks generated during pre-processing in Osprey. The ROI masks for the corticolimbic circuit were defined using Neurosynth (www.neurosynth.org/) term-based meta-analyses association maps using the search terms: ‘amygdala’, ‘hippocampus’, ‘acc’ and ‘nucleus accumbens’ (Figure 2A). FSL 6.0.7.17 was then used to threshold, lateralise and binarise these images.

#### Statistical analysis

#### Demographic and clinical data

Group comparisons of clinical and demographic data were performed in IBM SPSS statistics (v31.0.0). For the three groups, differences were assessed using one-way analysis of variance (ANOVA) for continuous variables and chi-squared tests for categorical variables. *Post-hoc* analyses were corrected for multiple comparisons using Tukey-HSD. Where applicable, group differences between the CHR-P and HC groups were tested using t-tests for continuous variables and chi-squared tests for categorical variables. Chlorpromazine equivalent antipsychotic doses were calculated using the chlorpromazineR web application (http://eebc.ca/chlorpromazineR_shiny/).

#### Combined rs-fMRI and ^1^H-MRS data analysis

All first- and second-level rs-fMRI analyses were performed within CONN. Our primary analyses investigated associations between regional Glx levels in the ACC and hippocampus and FC using ROI-to-ROI (and complementary seed-to-whole-brain) analyses. ROI connectivity matrices were estimated characterizing FC between each pair of regions among six ROIs for the hippocampus analyses, and seven ROIs for the ACC analyses. FC strength was represented by Fisher-transformed bivariate correlation coefficients from a general linear model (GLM), estimated separately for each pair of ROIs, characterizing the association between their BOLD signal timeseries. Data were entered into 2nd-level analyses and tested for Glx-FC associations per group and group interaction effects (how Glx-FC associations differ between HC and CHR-P), with age and sex as covariates. ROI-to-ROI results were thresholded at p < 0.05 for connections, with cluster-level significance p-false discovery rate (FDR) < 0.05. For completeness, seed-to-whole-brain connectivity maps were estimated for the Glx-FC associations and group interaction effects for the hippocampus and ACC (supplementary methods 1.4).

#### Individual modality fMRI and ^1^H-MRS data analysis

We also examined whether Glx levels in the two MRS voxels were different between groups using a repeated measures ANCOVA with group (HC, CHR-P and FEP) as a between-subject factor and region (ACC, hippocampus) as a within-subject factor. Based on the known association between ^1^H-MRS-derived Glx levels and age [4,39], we included age as a covariate and further examined associations between age and metabolite levels using two-tailed Pearson’s correlations. Significance for all analyses was determined at p < 0.05 with Bonferroni correction for *post hoc* multiple comparisons. We also examined group differences in FC from the MRS voxel seeds to the corticolimbic ROIs (and to the whole brain for completeness) by performing between-group analyses in each direction (HC>CHR-P; CHR-P>HC).

#### Exploratory analyses

Finally, as exploratory analyses, we measured the correlation between ACC and hippocampal Glx levels and symptom scores (CAARMS positive and negative, PANSS, HAM-A, HAM-D and WAIS-III) for HC, CHR-P, and FEP groups. We also assessed the correlation between the FC values for the Glx-FC group interactions and the above symptom scores. These statistical analyses were implemented in Python (version 3.12.2) using pandas for data handling and SciPy [40] for correlation analyses. As these were exploratory analyses, results were reported at an uncorrected level of p < 0.05.

## Results

### Sample Characteristics and Demographics

Participant demographic and clinical data are summarised in Table 1. After quality control, data from 23 HC, 22 CHR-P, and 10 FEP individuals were included. Separate tables for the participants included in each of the MRS-fMRI analyses (ACC and left hippocampus) are included in the supplementary materials (ST4 and ST5). There were no significant differences between groups in age (F=3.06 p=0.056) or sex (χ^2^=3.49, p=0.175), but there were significant differences in ethnicity (χ^2^=27.26, p<0.001) due to a significantly higher proportion of Asian individuals in the HC group compared to the CHR-P and FEP groups. There were also significant differences in HAM-A (F=18.8 p<0.001) and HAM-D (F=25.6 p<0.001) scores. *Post-hoc* comparison with Tukey-HSD correction showed higher depression and anxiety scores in CHR-P and FEP compared to HC (all corrected-p < 0.05).

**Table 1.**
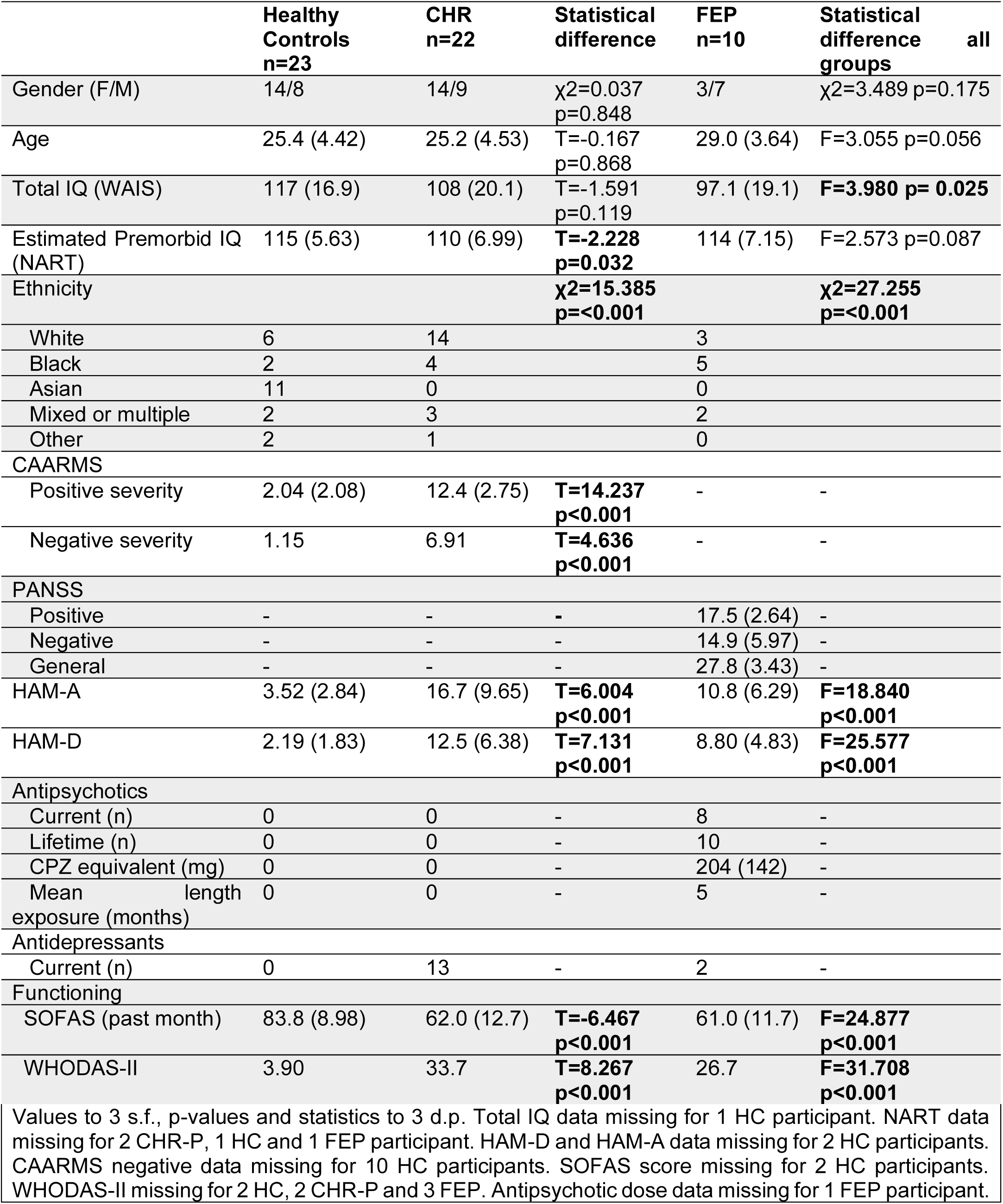
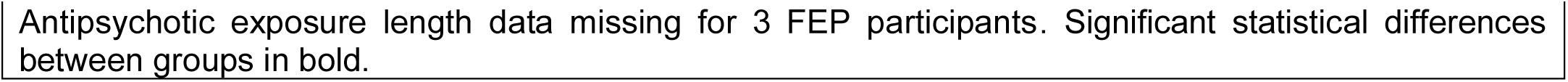
Demographic and Clinical Characteristics for study participants.

|  | Healthy Controls<br>n=23 | CHR<br>n=22 | Statistical<br>difference | FEP<br>n=10 | Statistical<br>difference<br>all<br>groups |
| --- | --- | --- | --- | --- | --- |
| Gender (F/M) | 14/8 | 14/9 | $\chi^2=0.037$<br>$p=0.848$ | 3/7 | $\chi^2=3.489$ $p=0.175$ |
| Age | 25.4 (4.42) | 25.2 (4.53) | $T=-0.167$<br>$p=0.868$ | 29.0 (3.64) | $F=3.055$ $p=0.056$ |
| Total IQ (WAIS) | 117 (16.9) | 108 (20.1) | $T=-1.591$<br>$p=0.119$ | 97.1 (19.1) | **$F=3.980$ $p=0.025$** |
| Estimated Premorbid IQ (NART) | 115 (5.63) | 110 (6.99) | **$T=-2.228$**<br>**$p=0.032$** | 114 (7.15) | $F=2.573$ $p=0.087$ |
| Ethnicity |  |  | **$\chi^2=15.385$**<br>**$p<0.001$** |  | **$\chi^2=27.255$**<br>**$p<0.001$** |
| White | 6 | 14 |  | 3 |  |
| Black | 2 | 4 |  | 5 |  |
| Asian | 11 | 0 |  | 0 |  |
| Mixed or multiple | 2 | 3 |  | 2 |  |
| Other | 2 | 1 |  | 0 |  |
| **CAARMS** |  |  |  |  |  |
| Positive severity | 2.04 (2.08) | 12.4 (2.75) | **$T=14.237$**<br>**$p<0.001$** | - | - |
| Negative severity | 1.15 | 6.91 | **$T=4.636$**<br>**$p<0.001$** | - | - |
| **PANSS** |  |  |  |  |  |
| Positive | - | - | - | 17.5 (2.64) | - |
| Negative | - | - | - | 14.9 (5.97) | - |
| General | - | - | - | 27.8 (3.43) | - |
| HAM-A | 3.52 (2.84) | 16.7 (9.65) | **$T=6.004$**<br>**$p<0.001$** | 10.8 (6.29) | **$F=18.840$**<br>**$p<0.001$** |
| HAM-D | 2.19 (1.83) | 12.5 (6.38) | **$T=7.131$**<br>**$p<0.001$** | 8.80 (4.83) | **$F=25.577$**<br>**$p<0.001$** |
| **Antipsychotics** |  |  |  |  |  |
| Current (n) | 0 | 0 | - | 8 | - |
| Lifetime (n) | 0 | 0 | - | 10 | - |
| CPZ equivalent (mg) | 0 | 0 | - | 204 (142) | - |
| Mean length exposure (months) | 0 | 0 | - | 5 | - |
| **Antidepressants** |  |  |  |  |  |
| Current (n) | 0 | 13 | - | 2 | - |
| **Functioning** |  |  |  |  |  |
| SOFAS (past month) | 83.8 (8.98) | 62.0 (12.7) | **$T=-6.467$**<br>**$p<0.001$** | 61.0 (11.7) | **$F=24.877$**<br>**$p<0.001$** |
| WHODAS-II | 3.90 | 33.7 | **$T=8.267$**<br>**$p<0.001$** | 26.7 | **$F=31.708$**<br>**$p<0.001$** |

Values to 3 s.f., p-values and statistics to 3 d.p. Total IQ data missing for 1 HC participant. NART data missing for 2 CHR-P, 1 HC and 1 FEP participant. HAM-D and HAM-A data missing for 2 HC participants. CAARMS negative data missing for 10 HC participants. SOFAS score missing for 2 HC participants. WHODAS-II missing for 2 HC, 2 CHR-P and 3 FEP. Antipsychotic dose data missing for 1 FEP participant.
Antipsychotic exposure length data missing for 3 FEP participants. Significant statistical differences between groups in bold.

### Interactions between Group, Glx and functional connectivity

For the ROI-to-ROI analyses there were significant group (CHR-P vs HC) x ACC Glx interactions on FC between the left NAc and the bilateral amygdala, right NAc and left amygdala, and bilateral NAc and bilateral hippocampus (F(2,39)=5.63, p-FDR=0.021) (Figure 2B and 2C). In CHR-P, there was a negative association between ACC Glx and FC between the left NAc and the bilateral hippocampus and amygdala, between the right NAc and bilateral hippocampus, as well as between the ACC and left hippocampus (F(2,39)=7.62, p-FDR=0.0048) (Figure 2D). These associations were not evident in HC (Figure 2E). Complementary seed-to-whole-brain analysis revealed further significant negative associations between ACC Glx levels and FC to left middle temporal gyrus (T(40)=-4.96, pFDR=0.009, Figure 3A and ST4) in CHR-P individuals, which were absent in HC.

There were no significant group x hippocampal Glx interactions on FC between our ROIs. Complementary seed-to-whole-brain analysis revealed, in the CHR-P group, a significant negative association between left hippocampus Glx levels and FC to the left parahippocampal gyrus and temporal fusiform cortex (T(40)=-4.96, pFDR=0.0078, Figure 3B and ST5), which was not significant in HC.

### Imaging results by modality

#### Glx levels in ACC and hippocampus

MRS spectral data quality and voxel tissue composition are summarised in Table 2. Supplementary materials (ST3) show these measures for the samples included in the MRS-fMRI analyses. Spectral and voxel overlap is presented in Figure 1 (and spectral fitting examples in Figure S1).

**Figure 1.**
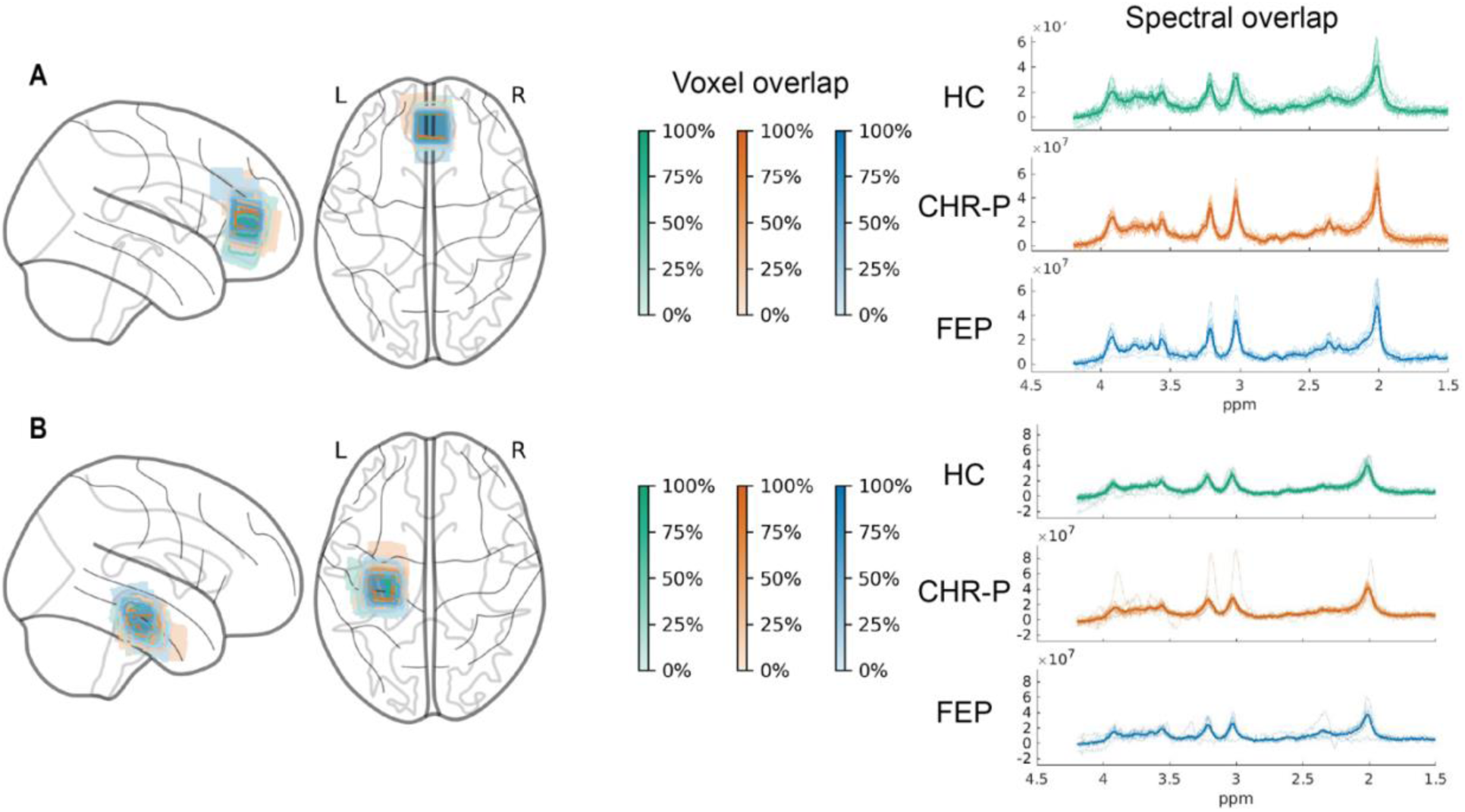
^1^H-MRS voxel placement overlap and spectral overlap. (A) Anterior cingulate cortex (ACC) and (B) Hippocampus normalised voxel placement and spectral overlap split by group. The colour of the voxel boxes on the glass brain indicates group and shading of the voxel lines indicates the percentage overlap across participants within the group. Fitted spectra for individual participants are plotted for each group. Green, healthy controls (HC); blue, clinical high risk for psychosis (CHR-P); orange, first episode psychosis (FEP). Voxel overlap images generated using an adapted version of MRSvoxel-plot with nilearn. ppm= parts per million.

**Figure 2.**
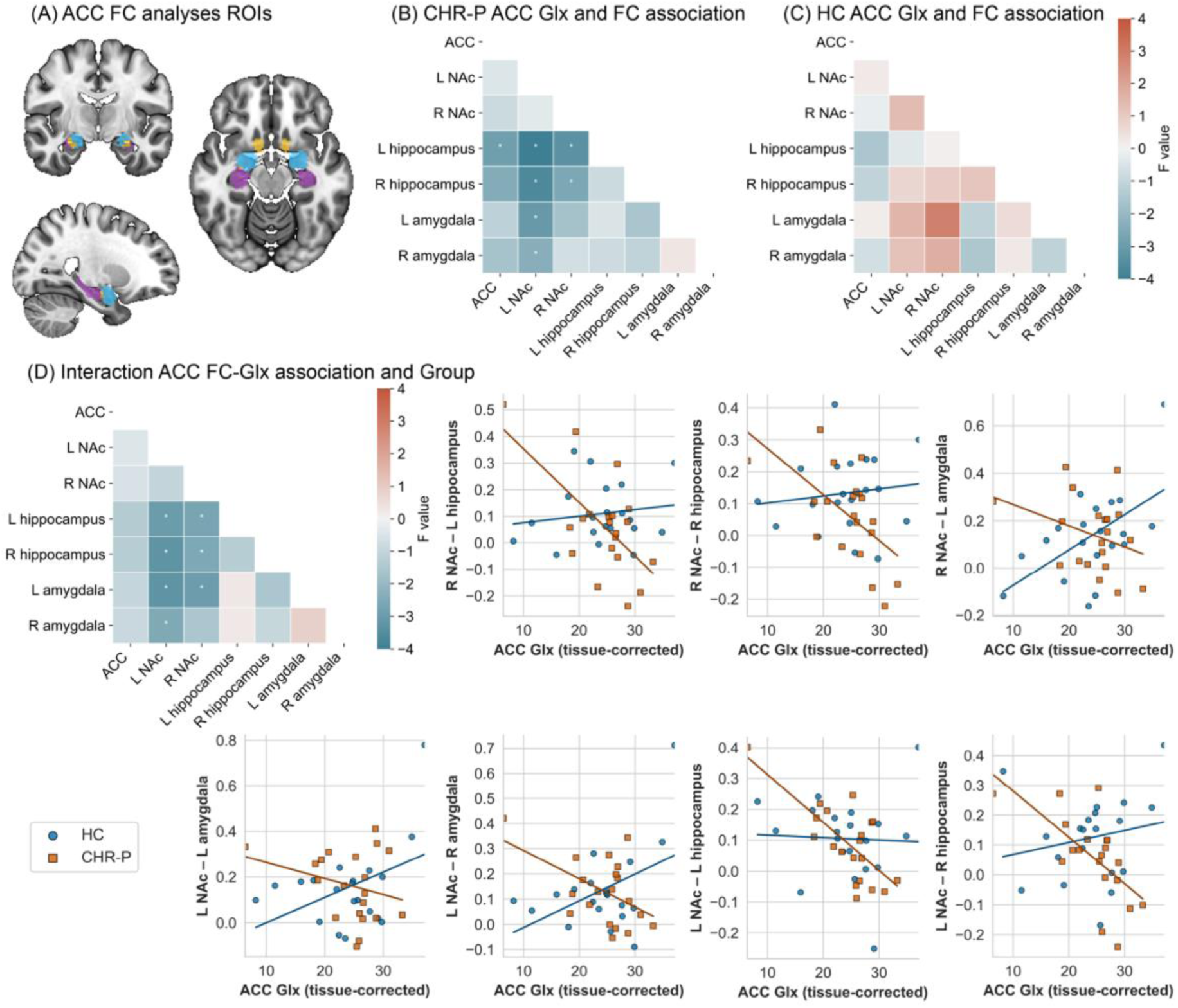
Region-of-interest (ROI) analyses of the association between anterior cingulate cortex (ACC) ^1^H-MRS Glx levels and resting-state functional connectivity compared by group: healthy controls (HC) and clinical high risk for psychosis (CHR-P). (A) Regions used for the ACC ROI-to-ROI FC analyses as defined using Neurosynth (www.neurosynth.org/). Amygdala= blue, hippocampus= purple, nucleus accumbens (NAc)= yellow. (B) F-value results for the association of CHR-P ACC Glx and ROI-to-ROI FC. Asterix indicated the connections where the association was significant after FDR correction (C) F-value plot for the association of HC ACC Glx and ROI-to-ROI FC. There were no ROI-to-ROI connections that showed a significant association with ACC Glx for the HC group. (D) F-value results for interaction of ACC Glx associations with FC and group (CHR-P > HC) for all ROI-to-ROI pairs. Asterix indicates the connections where the association was significantly different between groups after FDR correction. Scatter plots of the association between ACC Glx levels and functional connectivity between ROIs (y axis represents beta values). Plotted only for ROI-to-ROI pairs which had a significant group x Glx interaction. HC= blue, CHR-P= orange. L= Left, R= Right.

**Figure 3.**
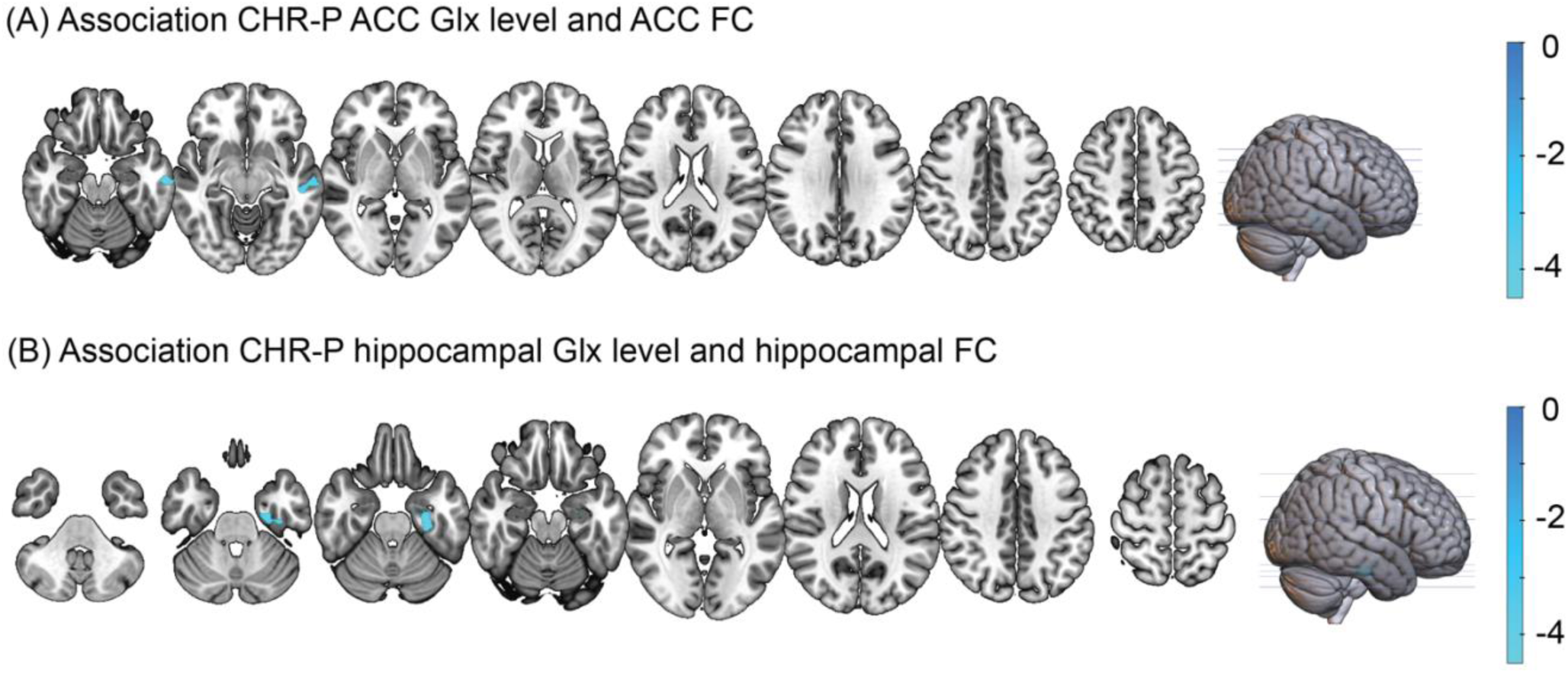
Seed-to-whole-brain analysis of the association between anterior cingulate cortex (ACC) and left hippocampal functional connectivity and ^1^H-MRS Glx. (A) Association CHR-P ACC Glx levels and ACC functional connectivity. (B) Association CHR-P hippocampal Glx levels and hippocampal functional connectivity. Blue colours indicate negative T-values.

Repeated-measures ANCOVA with age as a covariate revealed significant effects of group (F(2,43)=4.63, p=0.015) and voxel (F(1, 43)=13.78, p<0.001), but no significant voxel x group interaction (F(2,43)=0.370, p=0.693). Bonferroni-adjusted *post-hoc* analyses revealed that FEP individuals had significantly higher Glx than HC across both voxels (mean difference=4.38, SE=1.44, p=0.012). There was no significant main effect of age (F(1,43)=2.57, p=0.116). We observed a significant voxel x age interaction (F(2,43)=4.85, p=0.033) (Figure 4 A-C) and group x age interaction (F(3,43) =3.29, p=0.030), indicating the association between age and Glx differed between the groups and voxel locations, but follow-up pairwise comparisons were not statistically significant. Indeed, there was no significant correlation between age and ACC Glx levels (r=-0.26, n=47, p=0.077) or hippocampal Glx levels (r=0.20, n=47, p=0.176) across the groups, or within the individual groups (p>0.05, see Figure S2). Creatine FWHM was included as a covariate to account for potential impacts of spectral linewidth (a key measure of MRS spectral quality), and did not show a significant main effect (F=2.23, p=0.142) or interaction effects (p>0.05). This indicates that group differences in Glx levels are not significantly driven by differences in spectral linewidth between the groups. Sensitivity analyses for potential outliers did not change these results (supplementary results 2.2).

**Figure 4.**
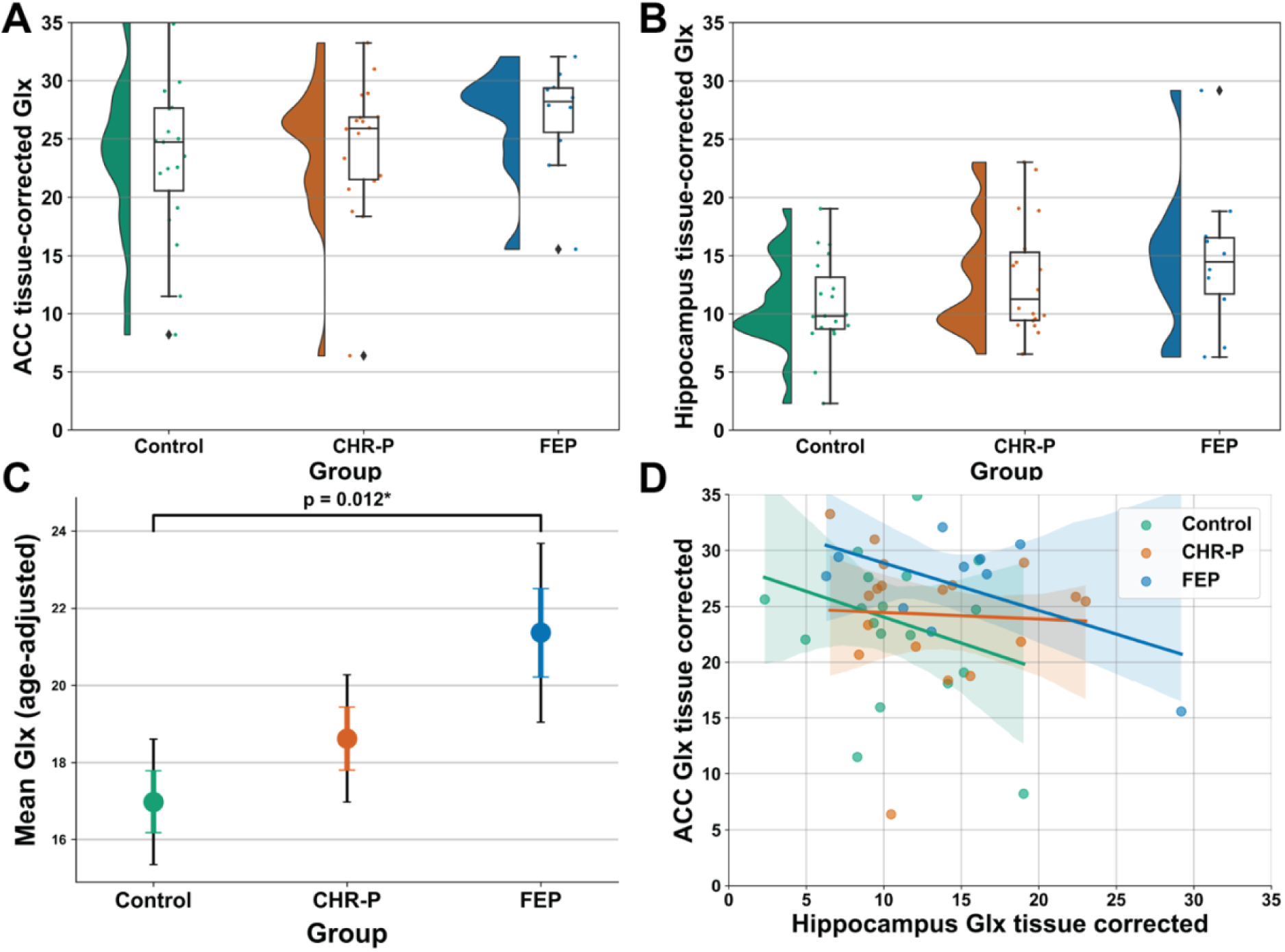
^1^H-MRS glutamate + glutamine (Glx) levels by group (healthy controls, clinical high risk for psychosis (CHR-P) and first episode psychosis (FEP)) and voxel region (hippocampus and anterior cingulate cortex (ACC)). Raincloud plot of tissue-corrected and water-scaled Glx levels for ACC (A) and hippocampus (B) split by groups. Showing median, interquartile range and spread of values. Diamonds represent outliers (>1.5 interquartile range). (C) Age-adjusted estimated marginal means from repeated measures ANCOVA for mean Glx split by group. Coloured bars indicate standard error, and black lines indicate 95% confidence intervals. *p-value from ANCOVA post-hoc pairwise comparisons with Bonferroni correction. (D) Scatter plot with regression lines split by group for Glx in the ACC and hippocampus. Shaded areas around regression lines represent 95% confidence intervals.

Supplementary analyses examined associations between Glx levels in the two regions using two-tailed Pearson’s correlations and age-adjusted partial correlations. There was no significant correlation (r=-0.164, n=47, p=0.270) or age-adjusted partial correlation (r=-0.119, n=47, p=0.433) between hippocampal and ACC Glx for the entire sample or within individual groups (p>0.05) (Figure 4D).

#### Functional connectivity

In the ROI-to-ROI and seed-to-whole-brain FC analyses, for both the ACC and the hippocampus, group contrasts were not significant (HC>CHR or CHR>HC), see supplementary materials for within-group FC results (supplementary results 2.3, Figure S3 and S4).

### Exploratory analyses

There were no significant correlations between ACC or hippocampal Glx levels and symptom scores (CAARMS scores, PANSS scores, HAM-A, HAM-D, WAIS-III, Trail making A-B) or functioning (SOFAS and WHODAS-II) in any of the three groups (all p>0.05, ST6 and ST7). In the FEP group, there was no significant correlation between antipsychotic dose (chlorpromazine equivalents) and Glx in the ACC (r=0.484, p=0.271) or hippocampus (r=0.059, p=0.900). Correlation results for the FC values from the ACC Glx x group interaction are reported in supplementary materials (supplementary results 2.4, ST10 and ST11).

## Discussion

Our main findings were that in CHR-P individuals, ACC Glx was negatively associated with connectivity within corticolimbic circuits, associations that were not evident in HCs. Furthermore, FEP individuals exhibited higher ACC and hippocampal Glx than HCs after controlling for age, but no Glx differences were observed between CHR-P individuals and HCs. There was also no significant group difference between CHR-P and HCs for FC from the ACC or the hippocampus. Collectively, these results suggest that there is altered coupling between glutamatergic metabolites and corticolimbic FC in people at high risk of psychosis, even in the absence of group-level Glx or FC differences.

Our finding that ACC Glx in CHR-P was negatively associated with connectivity within corticolimbic circuits (ACC-left hippocampus, left NAc-bilateral hippocampus, left NAc-bilateral amygdala and right NAc-bilateral hippocampus), with significant group differences in these relationships and no Glx-FC associations in HCs, supports the idea that neurochemical-functional coupling is disrupted in corticolimbic circuits in CHR-P individuals. Furthermore, exploratory seed-to-whole-brain analyses showed that higher ACC and hippocampal Glx were both related to lower temporal lobe connectivity in CHR-P. Overall, our findings align with prior reports in participants with chronic and first episode psychosis that glutamatergic dysregulation can disrupt corticolimbic circuit function, even when group-level differences in glutamatergic metabolites are modest or absent [25], and suggests that these disruptions can be identified in psychosis risk states not confounded by antipsychotic exposure. This supports the view that network-level Glx-FC interactions may be more sensitive early indicators of psychosis vulnerability than regional glutamatergic concentrations. One possible explanation for this is that baseline Glx levels differ between CHR-P individuals due to varying degrees of homeostatic compensation within glutamatergic systems. However, the functional consequences of these variations are likely to manifest at the post-synaptic or network level, which may be better captured by FC measures.

The absence of significant Glx alterations in CHR-P is consistent with previous meta-analytic evidence reporting no convergent prefrontal or hippocampal Glx differences in CHR-P compared to controls [10]. Indeed, the CHR-P and FEP literature reports smaller effect sizes, less consistent findings and more region-specific glutamatergic alterations compared to those observed in chronic schizophrenia [4,9,10]. Early glutamatergic alterations may be specific to the subgroup within the CHR-P population that will later transition to psychosis. This is supported by evidence that glutamatergic abnormalities are associated with poorer outcomes in CHR-P [41]. Group-level analyses may therefore dilute these effects. We did observe Glx elevations across the ACC and hippocampus in the FEP group compared to HCs. A subset of prior studies reported elevations in ACC Glx/glutamate in never-medicated early psychosis [42] and treatment-resistant individuals [5,8]. In contrast with prior evidence suggesting that antipsychotic exposure is associated with decreases in glutamatergic metabolites [3,4], in this small FEP sample who were all currently or previously exposed to antipsychotics, Glx was not associated with antipsychotic dose, although this may be due to reduced power to detect such correlations within our FEP sample.

We found a significant interaction between age and group suggesting group differences in the Glx-age relationship. However, ACC and hippocampal Glx levels were not significantly correlated with age in the full sample or within individual groups, in contrast to previous meta-regression findings of larger glutamate/glutamine decreases with age in schizophrenia compared to controls [9]. Similarly, a more recent meta-analysis found negative associations between age and medial frontal cortex glutamate in patients with schizophrenia, FEP and controls but concluded that lower glutamate levels in patients may be due to antipsychotic exposure rather than accelerated age-related decline [4]. These observations imply that age-related impacts on glutamatergic metabolites in FEP are potentially confounded by medication and are not well captured in cross-sectional designs. Longitudinal designs will be important to track dynamic changes in the relationship between glutamatergic metabolites and FC across the course of psychosis.

Our previous [^11^C]Ro15-4513 PET findings from the same individuals showed no significant difference in binding at the GABA_A_ receptors containing the α5 subunit (GABA_A_Rα5) in the hippocampus between HC, CHR-P and FEP [26]. However, brain-wide and hippocampal GABA_A_Rα5 covariance patterns were decreased in CHR-P and FEP individuals compared to HC, suggesting GABAergic alterations in at-risk/early psychosis may involve network-level rather than regional dysfunction [26]. The present MRS-fMRI findings similarly suggest alterations on a network-level in the absence of localised Glx alterations, suggesting that dysregulation of excitation-inhibition balance in at-risk/early psychosis may manifest within distributed networks rather than through focal metabolic abnormalities. Further exploration of the relationship between glutamatergic and GABAergic metabolites, and the relationship between GABAergic metabolites and FC, would provide valuable insights into the neurochemical basis of network dysfunction in early psychosis. However, interpretation of neuroimaging-based GABAergic measures is complicated by the fact that many GABAergic interneurons synapse onto other interneurons, making it difficult to infer the net inhibitory or excitatory impact [43].

To our knowledge, this was the first study to examine relationships between ^1^H-MRS Glx and resting-state FC in CHR-P individuals, benefitting from inclusion of two ^1^H-MRS voxel locations. Nonetheless, several limitations should be considered. Firstly, Glx provides only a composite measure of glutamate + glutamine from intra- and extracellular pools and is also influenced by other cellular processes such as glucose metabolism and glutathione synthesis [44]. However, presynaptic glutamate and glutamate metabolism are closely associated [45] and MRS-measured glutamate has also been associated with markers of synaptic function [46]. Secondly, the location of the voxel within the ACC is likely to influence the MRS-fMRI relationships, given known task-positive and task-negative subdivisions within the ACC. While voxel placement was guided by consistent anatomical landmarks, imperfect voxel overlap across participants and groups may have introduced some variability in the measured relationships. Furthermore, glutamate and glutamine concentrations vary according to region of the ACC [47] and glutamate levels in different ACC regions have been shown to have different relationships with FC [48]. MRS cannot easily distinguish these segregations at 3T due to limitations on voxel size. Finally, sample sizes were modest due to the PET-MR study design, COVID-19 disruptions, and quality control losses across both modalities, which limited power for FEP analyses and precluded examination of Glx-FC associations in the FEP group. *A priori* power calculations showed that we had >80% power to detect a 20% difference between CHR-P and HC groups in ACC Glx, but after quality control we were underpowered for left hippocampal Glx (n=24 in each group required for 80% power to detect a 20% difference) [49]. Larger, well-powered studies are also needed to characterise Glx-FC-symptom associations more robustly.

## Conclusions

Our results indicate that glutamatergic metabolite levels in hippocampus and ACC are inversely related to corticolimbic FC in individuals at CHR-P, suggesting subtle, network-level disruption of the coupling between neurochemistry and functional connectivity in the psychosis risk state. Future studies with larger sample sizes and longitudinal designs will be important for better understanding how these interactions develop across the course of psychosis, and their relevance to clinical outcomes.

## Data Availability Statement

MRS and resting-state fMRI data has been uploaded to an open repository and will be made public upon publication. Other study data and code is available from the corresponding author on reasonable request.

## Author Contributions

GM, PM and FET designed the research. GM obtained the funding for the study. PBL, JJS, MS, SK, AK and NRL conducted the research. JD conducted neuroimaging data acquisition. AdM, TJS and PFP provided clinical oversight. BH, NV and JD supported participant recruitment. EAR provided neuroimaging data acquisition oversight. DL provided MRS analytic support. AG wrote the original draft. AG made all figures/tables. All authors critically revised the article. All authors approved the last version. Supervision was provided by GM and PD.

## Funding

This research was funded by the Wellcome Trust & The Royal Society [Sir Henry Dale Fellowship 202397/Z/16/Z to GM]. This manuscript represents independent research partly funded by the NIHR Maudsley Biomedical Research Centre at South London and Maudsley NHS Foundation Trust and King’s College London. The views expressed are those of the author(s) and not necessarily those of the NIHR or the Department of Health and Social Care. AG was funded by the Wellcome Trust [223486/Z/21/Z to AG]. AAG received funding from USPHS NIMH MH57440. MV is supported by EU funding within the MUR PNRR “National Center for HPC, BIG DATA AND QUANTUMCOMPUTING (Project no. CN00000013 CN1), the Ministry of University and Research within the Complementary National Plan PNC DIGITAL LIFELONG PREVENTION – DARE (Project no PNC0000002_DARE), and by Fondo per il Programma Nazionale di Ricercae Progetti di Rilevante Interesse Nazionale (PRIN, Project no 2022RXM3H7).

For the purpose of open access, the author has applied a CC-BY public copyright licence to any Author Accepted Manuscript version arising from this submission.

## Competing Interests

GM has received consulting fees from Boehringer Ingelheim and speaker fees from Johnson & Johnson. PD has received speaker’s fees from Lundbeck and Janssen. GM is an editorial board member for Neuropsychopharmacology. AAG has received consulting fees from Alkermes, Lundbeck, Takeda, Roche, Lyra, Concert, Newron and SynAgile, and research funding from Lundbeck, Newron and Merck. EAR is a full-time employee of Perceptive Inc. (formally Invicro). MV is named as an inventor on a patent related to the use of dopaminergic imaging in mental Illness (ID: WO/2021/111116). The remaining authors have no conflicts of interest to declare.

## Notes

### Author Declarations

London/Surrey Research Ethics Committee gave approval for this work (17/LO/1130)

