## Supplementary Materials for "Associations between corticolimbic glutamatergic metabolites and functional connectivity in people at clinical high-risk for psychosis"

#### Title

**Corresponding author:**

Prof. Gemma Modinos

Department of Psychological Medicine

**Supplementary Figure 1- Sample spectra from Osprey for anterior cingulate cortex (ACC) and left hippocampus 1H-MRS PRESS for two sample participants. (A-B) Sample spectra**

### 1. Supplementary methodology

#### 1.1 MRI Acquisition

High-resolution T1-weighted anatomical images were acquired using an Inversion Recovery Fast Spoiled Gradient Echo (IR-SPGR) sequence (TE/TI/TR=2.996/400/6.992 ms; flip angle=11°; matrix=256x256x200; voxel size=1.2x1.05x1.05 $mm^3$).

Resting-state fMRI data was acquired with gradient-echo echo-planar imaging (EPI) using the following parameters: TE/TR= 30/2002ms; interleaved ascending acquisition; 35 contiguous slices; voxel size= 3x3x3.6 $mm^3$; flip angle 80°; 64x64 acquisition matrix; 300 volumes; total scan time 10 mins.

1H-MRS spectra were acquired from two voxels during the same scanning session: ACC (20x20x20 $mm^3$) and left hippocampus (20×20×15 $mm^3$). The ACC voxel was specified with the centre of the voxel placed along the sagittal midline, 16 mm above the anterior portion of the genu of the corpus callosum, 90° to the anterior commissure to posterior commissure line. Spectral data was acquired using point-resolved spectroscopy (PRESS) (TE/TR= 30/3000 ms; flip angle= 90°; 96 acquisitions with an 8-step phase cycle, spectral width= 5000 Hz, spectral data points= 4096 points). Water suppression was performed with the standard GE PROBE (proton brain examination) sequence, which implements standard chemically selective suppression (CHESS) water suppression pulses. For each spectrum, unsuppressed water reference spectra were acquired (16 transients) to perform eddy current correction and water scaling during spectral analysis.

#### 1.3 MRS methodology

##### *1H-MRS preprocessing*

MRS data was pre-processed and analysed using Osprey version 2.9.0 within Matlab R2024b. The preprocessing pipeline followed Osprey's standard procedures for raw data processing and preparation for quantification and modelling (Oeltzschner et al., 2020). Robust spectral registration was used to align individual transients. This approach applies spectral alignment with water and lipid removal. Eddy-current correction, spectral averaging and coil combination were performed using the water reference spectrum. Spectra were modelled using Osprey's built-in linear combination algorithm using the following modelling parameters: fit range 0.4-4ppm for metabolite spectra and 2.0-7.4 for water spectra; spline baseline modelling with knot spacing of 0.4 ppm. A standard basis set including 18 metabolites and 9 macromolecules/lipid components was selected to match the vendor, field strength, sequence type and echo time.

##### *Co-registration*

Structural T1 images were used to spatially map the voxel location onto each participant's native anatomical space and to extract voxel-specific tissue fractions for gray matter, white matter and cerebral spinal fluid. Co-registration was performed using Osprey's built in coregistration module and segmentation was performed using SPM12 (which is integrated into Osprey).

The hippocampus PRESS sequence was performed at the end of a 1 hour and 48-minute PET/MRI scanning session. This means there was almost 1 hour between the structural T1 image and this sequence. The final volume of the n-back fMRI task, which was performed immediately before the hippocampus PRESS sequence, was therefore used to assist with co-registration. Skull stripping was applied to the T1 image using FSL's brain extraction tool (BET) and the final volume of the functional sequence was extracted and upsampled to 1 mm isotropic resolution using FSL's FLIRT tool. The upsampled functional volume was registered to the skull-stripped T1 image using FLIRT producing a linear transformation matrix. This transformation matrix was inverted and applied to the T1 image to align it to the functional space.

##### *MRS-in-MRS checklist*

| 1. Hardware | |
|---|---|
| a. Field strength [T] | 3 T |
| b. Manufacturer | General Electric |
| c. Model (software version if available) | Signa simultaneous PET-MR |
| d. RF coils: nuclei (transmit/receive), number of channels, type, body part | 12-channel head coil |
| e. Additional hardware | N/A |
| 2. Acquisition | |

| | |
|---|---|
| a. Pulse sequence | Point-resolved spectroscopy (PRESS) |
| b. Volume of Interest (VOI) locations | Anterior cingulate cortex (ACC)<br><br>Left Hippocampus |
| c. Nominal VOI size [$cm^3$, $mm^3$] | ACC: 20 x 20 x 20 $mm^3$<br><br>Left hippocampus: 20 × 20 × 15 $mm^3$ |
| d. Repetition Time (TR), Echo Time (TE) [ms, s] | TR 3000 ms, TE 30 ms |
| e. Total number of Excitations or acquisitions per spectrum<br><br>In time series for kinetic studies<br><br>i. Number of Averaged spectra (NA) per time-point<br>ii. Averaging method (e.g. block-wise or moving average)<br>iii. Total number of spectra (acquired / in time-series) | 96 acquisitions with a 8-step phase cycle. For the unsuppressed water, 16 acquisitions with an 8-step phase cycle. |
| f. Additional sequence parameters (spectral width in Hz, number of spectral points, frequency offsets)<br><br>If STEAM:, Mixing Time (TM)<br><br>If MRSI: 2D or 3D, FOV in all directions, matrix size, acceleration factors, sampling method | 5000 Hz, 4096 points, -2 ppm frequency offset |
| g. Water Suppression Method | Chemically selective suppression (CHESS) |
| h. Shimming Method, reference peak, and thresholds for “acceptance of shim” chosen | Vendor (GE) automated B0 shimming |
| i. Triggering or motion correction method<br><br>(respiratory, peripheral, cardiac triggering, incl. device used and delays) | N/A |
| 3. Data analysis methods and outputs | |
| a. Analysis software | Osprey version 2.9.0 |
| b. Processing steps deviating from quoted reference or product | None |
| c. Output measure<br><br>(e.g. absolute concentration, institutional units, ratio) | Water-scaled tissue-corrected in molar units (mmol/kg) |
| d. Quantification references and assumptions, fitting model assumptions | Osprey’s built-in linear combination algorithm using the following modelling parameters: fit range 0.4-4ppm for metabolite spectra and 2.0-7.4 for water spectra; spline baseline modelling with knot spacing of 0.4 ppm. |

| | |
|---|---|
| | A standard basis set including 18 metabolites and 9 macromolecules/lipid components was selected to match the vendor, field strength, sequence type and echo time.<br><br>The basis set included simulated signals for ascorbate (Asc), aspartate (Asp), creatine (Cr, including creatine methylene; $CrCH_2$), gamma-aminobutyric acid (GABA), glycerophosphocholine (GPC), glutathione (GSH), glutamine (Gln), glutamate (Glu), myo-inositol (mI), lactate (Lac), N-acetylaspartate (NAA), N-acetylaspartylglutamate (NAAG), phosphocholine (PCh), phosphocreatine (PCr), phosphoethanolamine (PE), scyllo-inositol (sI), and taurine (Tau), as well as parameterized macromolecule components centered at approximately 0.9, 1.2, 1.4, 1.7 and 2.0 ppm (MM09, MM12, MM14, MM17, MM20) and lipid components centered at approximately 0.9, 1.3 and 2.0 ppm (Lip09, Lip13, Lip20)." |
| **4. Data Quality** | |
| a. Reported variables<br><br>(SNR, Linewidth (with reference peaks)) | See Table 2 for full quality measures and tissue composition values split by groups.<br><br>Mean ±SD values for whole sample after quality control:<br><br>ACC creatine SNR= 25.008 ± 6.445<br>ACC creatine FWHM= 8.520 ± 3.157<br>Hippocampus creatine SNR= 17.421 ± 3.246<br>Hippocampus creatine FWHM= 10.476 ± 2.873 |
| b. Data exclusion criteria | FWHM > 2SD; S/N < 2SD; visual inspection of spectra and voxel location |
| c. Quality measures of postprocessing Model fitting (e.g. CRLB, goodness of fit, SD of residual) | Relative fit quality (ratio of the fit residual amplitude to the standard deviation of the noise). See Table 2 for full values split by group.<br><br>Mean ± SD values for whole sample after quality control:<br><br>ACC = 1.375 ± 0.267<br><br>Hippocampus= 1.263 ± 0.227 |
| d. Sample Spectrum | Figure 1 and supplementary figure 1 |

#### 1.3 fMRI methodology

##### *fMRI preprocessing*

The rs-fMRI data was preprocessed using fMRIPrep 25.0.0 (Esteban et al., 2019) and CONN 22.v2507 (Whitfield-Gabrieli & Nieto-Castanon, 2012). Structural T1 images were corrected for intensity non-uniformity using N4 bias field correction, skull-stripped, tissue segmented and normalised to MNI space. The rs-fMRI data underwent reference volume estimation and co-registration to the anatomical image. Participants were excluded if they had a mean framewise displacement (FD) > 0.5 mm. After importing into CONN, the functional data was spatially smoothed using 6 mm Gaussian kernel and denoised using a standard denoising pipeline including removal of white matter and CSF signal using the first five components of aCompCor, regression of motion parameters and their first-order derivatives, despiking, linear de-trending, scrubbing and band-pass filtering (0.008-0.09 Hz). Further details on preprocessing are provided in fMRIprep and CONN boiler plate outputs below. All first- and second-level fMRI analyses were performed within CONN.

##### *fMRI prep boiler plate output*

Results included in this manuscript come from preprocessing
performed using *fMRIPrep* 25.0.0
(@fmriprep1; @fmriprep2; RRID:SCR_016216),
which is based on *Nipype* 1.9.2
(@nipype1; @nipype2; RRID:SCR_002502).

Anatomical data preprocessing

: A total of 1 T1-weighted (T1w) images were found within the input
BIDS dataset. The T1w image was corrected for intensity
non-uniformity (INU) with `N4BiasFieldCorrection` [@n4], distributed with ANTs 2.5.4
[@ants, RRID:SCR_004757], and used as T1w-reference throughout the workflow.
The T1w-reference was then skull-stripped with a *Nipype* implementation of
the `antsBrainExtraction.sh` workflow (from ANTs), using OASIS30ANTs
as target template.
Brain tissue segmentation of cerebrospinal fluid (CSF),
white-matter (WM) and gray-matter (GM) was performed on
the brain-extracted T1w using `fast` [FSL (version unknown), RRID:SCR_002823, @fsl_fast].
Volume-based spatial normalization to one standard space (MNI152NLin2009cAsym) was performed through
nonlinear registration with `antsRegistration` (ANTs 2.5.4),
using brain-extracted versions of both T1w reference and the T1w template.

The following template was were selected for spatial normalization
and accessed with *TemplateFlow* [24.2.2, @templateflow]:
*ICBM 152 Nonlinear Asymmetrical template version 2009c* [@mni152nlin2009casym, RRID:SCR_008796; TemplateFlow ID: MNI152NLin2009cAsym].

Functional data preprocessing

: For each of the 1 BOLD runs found per subject (across all
tasks and sessions), the following preprocessing was performed.
First, a reference volume was generated,
using a custom methodology of *fMRIPrep*, for use in head motion correction.
Head-motion parameters with respect to the BOLD reference
(transformation matrices, and six corresponding rotation and translation
parameters) are estimated before any spatiotemporal filtering using
`mcflirt` [FSL <ver>, @mcflirt].
The BOLD reference was then co-registered to the T1w reference using
`mri_coreg` (FreeSurfer) followed by `flirt` [FSL <ver>, @flirt]
with the boundary-based registration [@bbr] cost-function.
Co-registration was configured with six degrees of freedom.
Several confounding time-series were calculated based on the
*preprocessed BOLD*: framewise displacement (FD), DVARS and
three region-wise global signals.
FD was computed using two formulations following Power (absolute sum of
relative motions, @power_fd_dvars) and Jenkinson (relative root mean square
displacement between affines, @mcflirt).
FD and DVARS are calculated for each functional run, both using their
implementations in *Nipype* [following the definitions by @power_fd_dvars].
The three global signals are extracted within the CSF, the WM, and
the whole-brain masks.
Additionally, a set of physiological regressors were extracted to
allow for component-based noise correction [*CompCor*, @compcor].
Principal components are estimated after high-pass filtering the
*preprocessed BOLD* time-series (using a discrete cosine filter with
128s cut-off) for the two *CompCor* variants: temporal (tCompCor)
and anatomical (aCompCor).
tCompCor components are then calculated from the top 2% variable
voxels within the brain mask.
For aCompCor, three probabilistic masks (CSF, WM and combined CSF+WM)
are generated in anatomical space.
The implementation differs from that of Behzadi et al. in that instead
of eroding the masks by 2 pixels on BOLD space, a mask of pixels that
likely contain a volume fraction of GM is subtracted from the aCompCor masks.
This mask is obtained by thresholding the corresponding partial volume map at 0.05, and it ensures components are not extracted
from voxels containing a minimal fraction of GM.
Finally, these masks are resampled into BOLD space and binarized by
thresholding at 0.99 (as in the original implementation).
Components are also calculated separately within the WM and CSF masks.
For each CompCor decomposition, the *k* components with the largest singular
values are retained, such that the retained components' time series are
sufficient to explain 50 percent of variance across the nuisance mask (CSF,
WM, combined, or temporal). The remaining components are dropped from

consideration.
The head-motion estimates calculated in the correction step were also placed within the corresponding confounds file.
The confound time series derived from head motion estimates and global signals were expanded with the inclusion of temporal derivatives and quadratic terms for each [@confounds_satterthwaite_2013].
Frames that exceeded a threshold of 0.5 mm FD or 1.5 standardized DVARS were annotated as motion outliers.
Additional nuisance timeseries are calculated by means of principal components analysis of the signal found within a thin band (*crown*) of voxels around the edge of the brain, as proposed by [@patriat_improved_2017].
All resamplings can be performed with *a single interpolation step* by composing all the pertinent transformations (i.e. head-motion transform matrices, susceptibility distortion correction when available, and co-registrations to anatomical and output spaces).
Gridded (volumetric) resamplings were performed using `nitransforms`, configured with cubic B-spline interpolation.

Many internal operations of *fMRIPrep* use *Nilearn* 0.11.1 [@nilearn, RRID:SCR_001362], mostly within the functional processing workflow.
For more details of the pipeline, see [the section corresponding to workflows in *fMRIPrep*'s documentation](https://fmriprep.readthedocs.io/en/latest/workflows.html "FMRIPrep's documentation").

##### Copyright Waiver

The above boilerplate text was automatically generated by fMRIPrep with the express intention that users should copy and paste this text into their manuscripts *unchanged*.
It is released under the [CC0](https://creativecommons.org/publicdomain/zero/1.0/) license.

#### *CONN boiler plate output*

Analyses of fMRI data were performed using CONN[1] (RRID:SCR_009550) release 22.v2407[2] and SPM[3] (RRID:SCR_007037) release 12.7771.

Preprocessing: Functional data were smoothed using spatial convolution with a Gaussian kernel of 6 mm full width half maximum (FWHM).

Denoising: In addition, functional data were denoised using a standard denoising pipeline[4] including the regression of potential confounding effects characterized by white matter timeseries (5 CompCor noise components), CSF timeseries (5 CompCor noise components), motion parameters and their first order derivatives (12 factors)[5], session effects and their first order derivatives (2 factors), scrubbing_noGSC_FD0_9 regressors (52 components), and linear trends (2 factors) within each functional run, followed by bandpass frequency filtering of the BOLD timeseries[6] between 0.008 Hz and 0.09 Hz. CompCor[7,8] noise components within white matter and CSF were estimated by computing the average BOLD signal as well as the largest principal components orthogonal to the BOLD average, motion parameters within each subject's eroded segmentation masks. From the number of noise terms included in this denoising strategy, the effective degrees of freedom of the BOLD signal after denoising were estimated to range from 72.8 to 89.9 (average 88) across all subjects[9].

First-level analysis: Seed-based connectivity maps (SBC) were estimated characterizing the spatial pattern of functional connectivity with a seed area. Seed regions included ACC_voxel. Functional connectivity strength was represented by Fisher-transformed bivariate correlation coefficients from a weighted general linear model (weighted-GLM[10]), estimated separately for each seed area and target voxel, modeling the association between their BOLD signal timeseries. In order to compensate for possible transient magnetization effects at the beginning of each run, individual scans were weighted by a step function convolved with an SPM canonical hemodynamic response function and rectified. Hippocampus: Seed-based connectivity maps (SBC) were estimated characterizing the spatial pattern of functional connectivity with a seed area. Seed regions included L-hippocampus_voxel. Functional connectivity strength was represented by Fisher-transformed bivariate correlation coefficients from a weighted general linear model (weighted-GLM[10]), estimated separately for each seed area and target voxel, modeling the association between their BOLD signal timeseries. In order to compensate for possible transient magnetization effects at the beginning of each run, individual scans were weighted by a step function convolved with an SPM canonical hemodynamic response function and rectified.

ACC_voxel_to_ROIs: ROI-to-ROI connectivity (RRC) matrices were estimated characterizing the functional connectivity between each pair of regions among 7 ROIs. Functional connectivity strength was represented by Fisher-transformed bivariate correlation coefficients from a general linear model (weighted-GLM[10]), estimated separately for each pair of ROIs, characterizing the association between their BOLD signal timeseries. In order to compensate for possible transient magnetization effects at the beginning of each run, individual scans were weighted by a step function convolved with an SPM canonical hemodynamic response function and rectified. HIPPO_voxel_to_ROIs: ROI-to-ROI connectivity (RRC) matrices were estimated characterizing the functional connectivity between each pair of regions among 6 ROIs. Functional connectivity strength was represented by Fisher-transformed bivariate correlation coefficients from a general linear model (weighted-GLM[10]), estimated separately for each pair of ROIs, characterizing the association between their BOLD signal timeseries. In order to compensate for possible transient magnetization effects at the beginning of each run, individual scans were weighted by a step function convolved with an SPM canonical hemodynamic response function and rectified.

Group-level analyses seed-to-voxel: were performed using a General Linear Model (GLM[11]). For each individual voxel a separate GLM was estimated, with first-level connectivity measures at this voxel as dependent variables (one independent sample per subject and one measurement per task or experimental condition, if applicable), and groups or other subject-level identifiers as independent variables. Voxel-level hypotheses were evaluated using multivariate parametric statistics with random-effects across subjects and sample covariance estimation across multiple measurements. Inferences were performed at the level of individual clusters (groups of contiguous voxels). Cluster-level inferences were based on parametric statistics from Gaussian Random Field theory[12,13]. Results were thresholded using a combination of a cluster-forming $p < 0.001$ voxel-level threshold, and a familywise corrected p-FDR < 0.05 cluster-size threshold[14].

Group-level analyses ROI-to-ROI: were performed using a General Linear Model (GLM[11]). For each individual connection a separate GLM was estimated, with first-level connectivity measures at this connection as dependent variables (one independent sample per subject and one measurement per task or experimental condition, if applicable), and groups or other subject-level identifiers as independent variables. Connection-level hypotheses were evaluated using multivariate parametric statistics with random-effects across subjects and sample covariance estimation across multiple measurements. Inferences were performed at the level of individual clusters (groups of similar connections). Cluster-level inferences were based on parametric statistics within- and between- each pair of networks (Functional Network Connectivity[12]), with networks identified using a completelinkage hierarchical clustering procedure[13] based on ROI-to-ROI anatomical proximity and functional similarity metrics[14]. Results were thresholded using a combination of a $p < 0.05$ connection-level threshold and a familywise corrected p-FDR < 0.05 cluster-level threshold[15].

#### 1.4 Combined fMRI and 1H-MRS data analyses

*Seed-to-whole-brain analyses*

FC strength was represented by Fisher-transformed bivariate correlation coefficients from a weighted GLM, estimated separately for each seed and target voxel. Data were entered into 2nd-level analyses and tested for Glx-FC associations per group and group interaction effects (how Glx-FC associations differ between HC and CHR-P) with age and sex as covariates. Seed-to-whole-brain results used a cluster-forming threshold of p<0.001 and p-FDR<0.05 for cluster size.

### 2. Supplementary Results

#### 2.1 MRS quality measures comparisons between groups

For the ACC Glx analyses there was a significant group difference for water (F=4.03, p=0.025) and creatine FWHM (F=6.35, p=0.004). *Post hoc* analyses revealed that this was driven by the CHR-P group having significantly lower creatine FWHM (mean difference=3.24, p=0.003) and water FWHM (mean difference= 2.33, p=0.019) compared to HCs. There was no significant difference for relative fit quality for the ACC (F=2.003, p=0.147) or the hippocampus (F=0.191, p=0.827). There was a significant difference in voxel tissue composition for gray matter (F=6.89, p=0.002) and CSF (F=4.06, p=0.024) in the hippocampus Glx analyses.

#### 2.2 Glx levels in ACC and hippocampus repeated measures ANCOVA-sensitivity analyses with removal of outliers

For the Glx repeated measures ANCOVA analyses we performed a sensitivity analyses excluding outliers using the interquartile range (IQR) method, whereby values lying more than 1.5 × IQR below the first quartile or above the third quartile for a given variable were excluded from analysis. The above findings remained significant (effect of group F(2,40)=4.13, p=0.023; interaction voxel x age F(1,40)=9.10, p=0.004).

#### 2.3 Resting-state functional connectivity within-group results

In the seed-to-voxel FC analyses, within each group (CHR-P and HC), the ACC showed significant positive FC with other regions of the cingulate cortex, the frontal medial cortex, frontal pole, paracingulate cortex, lateral occipital cortex and temporal cortex (p-FDR < 0.05; Supplementary Figures 4A and 4B). For the CHR-P group there was also significant FC with the bilateral hippocampus and the surrounding parahippocampal gyrus and temporal fusiform cortex (p-FDR < 0.05; Supplementary Figure 3B). Group contrasts were not significant (HC>CHR or CHR>HC).

For the hippocampus seed, seed-to-voxel analyses revealed significant positive associations between FC to large parts of the temporal cortex, frontal medial cortex, postcentral gyrus, amygdala and right hippocampus (p-FDR < 0.05; Supplementary Figure 4C and 4D) in CHR-P and HC groups. The CHR-P group also showed significant positive FC with cerebellar regions. There were no significant results for the group contrasts (HC>CHR or CHR>HC).

#### 2.4 Correlations between ACC Glx x group interaction FC beta values and symptoms

There was no significant correlation between the FC beta values for the ACC Glx x group interaction ROI-to-ROI analyses and symptoms score for the CHR-P group (all p>0.05, ST8). For the HC group, the beta values for the ACC Glx x group interaction negatively correlated with depression scores (HAM-D) for ROI-to-ROI connections from the right NAc to the left amygdala (r= -0.508, uncorrected-p=0.0265) and right amygdala (r=-0.520, uncorrected-p=0.0226) (ST9).

#### 3. Supplementary Tables

**Supplementary Table 1**- Sample sizes included in MRS and combined MRS-fMRI analyses after quality control

| | ACC | | ACC and Hippocampus | Hippocampus | | MRS combined or MRS /fMRI |
|---|---|---|---|---|---|---|
| Group (total n) | MRS (only) | fMRI and MRS | MRS | MRS (only) | fMRI and MRS | In any analyses |
| HC (24) | 20 | 20 | 19 | 22 | 22 | 22 |
| CHR (24) | 22 | 20 | 18 | 19 | 18 | 23 |
| FEP (10) | 10 | 7* | 10 | 10 | 7* | 10 |
| TOTAL (58) | 52 | 47 | 47 | 51 | 47 | 55 |

*Note that FEP group were not included in the fMRI and MRS combined analyses due to n=7 after quality control of both modalities.

**Supplementary Table 2-** Antipsychotic medication information for FEP

| | FEP (n=10) |
|---|---|
| Antipsychotics current (n) | 8 |
| Antipsychotics lifetime (n) | 10 |
| Antipsychotics | Olanzapine- 2<br>Risperidone- 2<br>Quetiapine- 1<br>Aripiprazole- 4<br>Unknown- 1 |
| Mean antipsychotics exposure (months) | 5* |
| Antipsychotic dose (CPZ equivalent mg) | 204.3** |

*Exposure length data missing for 3 FEP participants **Dose data missing for 1 FEP participant

**Supplementary Table 3-** Magnetic Resonance Spectroscopy (MRS) data quality and tissue composition for the anterior cingulate cortex (ACC) and left hippocampus voxels

| | **ACC Voxel** | | | | **Hippocampus Voxel** | | | |
|---|---|---|---|---|---|---|---|---|
| | **Healthy Controls n=19** | **CHR n=18** | **FEP n=10** | **Statistical difference** | **Healthy Controls n=19** | **CHR n=18** | **FEP n=10** | **Statistical difference** |
| Glx levels (mmol/kg) | 23.673 (7.147) | 24.270 (5.997 | 26.859 (4.786) | F=0.878 p=0.423 | 10.780 (4.022) | 13.087 (4.942) | 14.750 (6.475) | F=2.295 p=0.113 |
| Creatine SNR | 22.632 (6.448) | 27.005 (4.574) | 25.929 (8.295) | F=2.394 p=0.103 | 18.446 (3.452) | 16.205 (2.482) | 17.665 (3.625) | F=2.373 p=0.105 |
| Creatine FWHM (Hz) | 10.275 (3.321) | 7.030 (1.909) | 7.866 (3.255) | **F=6.352 p=0.004** | 9.810 (3.239) | 11.627 (2.578) | 9.667 (2.088) | F=2.503 p=0.093 |
| Water FWHM (Hz) | 9.285 (2.787) | 6.960 (1.327) | 8.347 (3.414) | **F=4.026 p=0.025** | 10.544 (2.982) | 11.853 (2.351) | 10.825 (2.564) | F=1.184 p=0.316 |
| Residual water amplitude | 8002076 (4023368.0) | 8534906 (3820560.7) | 9572197 (4295901.5) | F=0.504 p=0.608 | 11531026 (2765477.8) | 13933725 (7625400.7) | 9139012 (4910643.8) | F=2.509 p=0.093 |
| Frequency shift (Hz) | -2.592 (2.180) | -2.284 (0.917) | -1.767 (1.566) | F=0.805 p=0.454 | -1.302 (2.444) | -1.744 (3.200) | -2.754 (2.016) | F=0.956 p=0.392 |
| Relative fit quality | 1.467 (0.353) | 1.320 (0.192) | 1.299 (0.267) | F=2.003 p=0.147 | 1.239 (0.246) | 1.273 (0.215) | 1.290 (0.226) | F=0.191 p=0.827 |
| GM Fraction | 0.655 (0.044) | 0.627 (0.076) | 0.623 (0.052) | F=1.355 p=0.269 | 0.706 (0.070) | 0.562 (0.165) | 0.671 (0.102) | **F=6.890 p=0.002** |
| WM Fraction | 0.107 (0.041) | 0.105 (0.046) | 0.094 (0.054) | F=0.307 p=0.737 | 0.206 (0.109) | 0.246 (0.130) | 0.252 (0.123) | F=0.689 p=0.503 |
| CSF Fraction | 0.238 (0.054) | 0.267 (0.056) | 0.283 (0.063) | F=2.435 p=0.099 | 0.088 (0.077) | 0.192 (0.183) | 0.077 (0.054) | **F=4.063 p=0.024** |

Values and statistics to 3d.p apart from residual water amplitude.
Glx (glutamate + glutamine)- tissue-corrected water-scaled.
Residual water amplitude in arbitrary units.
Relative fit quality= ratio of the fit residual amplitude to the standard deviation of the noise. Values close to 1 indicate a perfect fit. Values <1 indicate over-fitting of the data. Values significantly >1 indicate significant differences between the fit and the data.
Significant statistical differences between groups in bold.
Numbers after quality control and including only participants with both ACC and hippocampus MRS data.

**Supplementary Table 4-** Demographics and Clinical Characteristics for ACC fMRI analyses

| | **Healthy Controls n=20** | **CHR n=20** | **Statistical difference** |
|---|---|---|---|
| Gender (F/M) | 14/6 | 12/8 | $\chi2$=0.440 p=0.507 |
| Age | 25.06 (4.59) | 25.24 (4.75) | T= 0.119 P=0.906 |
| Total IQ (WAIS) | 113.90 (16.00) | 109.30 (19.80) | T=-0.795 P= 0.432 |
| Estimated Premorbid IQ (NART) | 114.64 (5.71) | 110.68 (7.09) | **T=-1.898 P=0.066** |
| Ethnicity | | | **$\chi2$=17.600 p=0.001** |
| White | 3 | 12 | |
| Black | 2 | 4 | |
| Asian | 11 | 0 | |
| Mixed or multiple | 2 | 1 | |
| Other | 2 | 3 | |
| CAARMS | | | |
| Positive severity | 2.15 (2.08) | 12.55 (2.42) | **T=14.574 P= <0.001** |
| Negative severity | 1.15 (1.73) | 6.90 (4.28) | **T=4.586 P=<0.001** |
| HAM-A | 3.63 (2.97) | 16.70 (9.76) | **T=5.592 P=<0.001** |
| HAM-D | 2.11 (1.91) | 12.50 (6.68) | **T=6.525 P=<0.001** |
| Antidepressants | | | |
| Current (n) | 0 | 12 | |
| Functioning | | | |
| SOFAS (past month) | 83.68 (9.39) | 61.45 (12.93) | **T=-6.116 P=<0.001** |
| WHODAS-II | 4.11 (4.60) | 34.00 (16.28) | **T=7.494 P=<0.001** |

*Values to 3 s.f, p-values and statistics to 3 d.p*
*HAM-A: Hamilton Anxiety Rating Scale. HAM-D: Hamilton Depression Rating Scale. CAARMS: Comprehensive Assessment of At-Risk Mental States. SOFAS: Social and Occupational Functioning Assessment Scale. WHODAS-II: World Health Organization Disability Assessment Schedule. WAIS-III: Wechsler Adult Intelligence Scale, 3rd Edition. NART: National Adult Reading Test.*
*Total IQ data missing for 1 HC participant. NART data missing for 2 CHR-P and 1 HC participant. HAM-D and HAM-A data missing for 2 HC participants. CAARMS negative data missing for 10 HC participants. SOFAS score missing for 2 HC participants. WHODAS-II missing for 1 HC and 2 CHR-P.*

**Supplementary Table 5-** Demographics and Clinical Characteristics for hippocampus fMRI analyses

| | **Healthy Controls n=22** | **CHR n=18** | **Statistical difference** |
|---|---|---|---|
| Gender (F/M) | 13/9 | 11/7 | $\chi2$=0.017 p=0.897 |
| Age | 25.23 (4.43) | 25.95 (4.56) | T= 0.506 P=0.308 |
| Total IQ (WAIS) | 115.86 (16.74) | 106.06 (19.69) | T=-1.681 P=0.101 |
| Estimated Premorbid IQ (NART) | 114.45 (5.56) | 110.87 (6.80) | T=-1.764 P=0.086 |
| Ethnicity | | | **$\chi2$=14.141 p=0.007** |
| White | 6 | 12 | |
| Black | 2 | 3 | |
| Asian | 10 | 0 | |
| Mixed or multiple | 2 | 0 | |
| Other | 2 | 3 | |
| CAARMS | | | |
| Positive severity | 2.14 (2.08) | 12.94 (2.29) | **T=15.645 P<0.001** |
| Negative severity | 1.17 (1.80) | 7.67 (4.04) | **T= 5.211 P<0.001** |
| HAM-A | 3.55 (2.91) | 16.44 (8.47) | **T=6.407 P<0.001** |
| HAM-D | 2.05 (1.76) | 12.44 (6.17) | **T=7.228 P<0.001** |
| Antidepressants | | | |
| Current (n) | 0 | 11 | |
| Functioning | | | |
| SOFAS (past month) | 83.45 (9.10) | 62.11 (13.41) | **T=-5.791 P<0.001** |
| WHODAS-II | 3.90 (4.35) | 33.69 (16.65) | **T= 7.883 P<0.001** |

*Values to 3 s.f, p-values and statistics to 3 d.p*
*HAM-A: Hamilton Anxiety Rating Scale. HAM-D: Hamilton Depression Rating Scale. CAARMS: Comprehensive Assessment of At-Risk Mental States. SOFAS: Social and Occupational Functioning Assessment Scale. WHODAS-II: World Health Organization Disability Assessment Schedule. WAIS-III: Wechsler Adult Intelligence Scale, 3rd Edition. NART: National Adult Reading Test.*
*Total IQ data missing for 1 HC participant. NART data missing for 1 CHR-P and 1 HC participant. HAM-D and HAM-A data missing for 1 HC participant. CAARMS negative data missing for 7 HC participants. SOFAS score missing for 1 HC participant. WHODAS-II missing for 2 HC and 2 CHR-P.*

**Supplementary Table 6-** Quality measures for MRS and fMRI data included in the ACC and hippocampus analyses

| | **ACC Voxel** | | | **Hippocampus Voxel** | | |
|---|---|---|---|---|---|---|
| | **Healthy Controls n=20** | **CHR-P n=20** | **Statistical difference** | **Healthy Controls n=22** | **CHR-P n=18** | **Statistical difference** |
| Mean FD (mm) | 0.118 (0.032) | 0.146 (0.055) | T=1.923 P=0.062 | 0.118 (0.031) | 0.143 (0.055) | T=1.844 P=0.073 |
| Glx levels (mmol/kg) | 23.977 (7.088) | 24.442 (5.834) | T=0.227 P=0.822 | 11.251 (5.252) | 13.214 (4.943) | T=1.207 P=0.235 |
| Creatine SNR | 22.584 (6.280) | 26.551 (4.647) | **T=2.271 P=0.029** | 18.234 (3.396) | 16.6301 (3.350) | T=-1.494 P=0.143 |
| Creatine FWHM (Hz) | 10.250 (3.234) | 7.305 (2.367) | **T=-3.286 P=0.002** | 9.823 (3.040) | 11.466 (2.672) | T=1.794 P=0.081 |
| Water FWHM (Hz) | 9.236 (2.722) | 7.124 (1.356) | **T=-3.105 P=0.004** | 10.539 (2.775) | 11.696 (2.464) | T=1.378 P=0.176 |
| Residual water amplitude | 7845961.52 (3977807.38) | 8040300.61 (3688210.65) | T=0.160 P=0.874 | 11415865.06 (2705986.42) | 13591493.66 (7599212.03) | T=-1.252 P=0.218 |
| Frequency shift (Hz) | -2.615 (2.124) | -2.276 (0.924) | T=0.653 P=0.517 | -1.507 (2.444) | -1.688 (3.207) | T=-0.103 P=0.840 |
| GM Fraction | 0.653 (0.043) | 0.632 (0.072) | T=-1.132 P=0.265 | 0.706 (0.082) | 0.549 (0.158) | **T=-4.063 P<0.001** |
| WM Fraction | 0.104 (0.043) | 0.105 (0.042) | T=0.083 P=0.934 | 0.210 (0.120) | 0.262 (0.139) | T=1.276 P=0.210 |
| CSF Fraction | 0.243 (0.057) | 0.263 (0.054) | T=1.142 P=0.261 | 0.084 (0.073) | 0.189 (0.185) | **T=2.437 P=0.020** |

*Values and statistics to 3d.p apart from residual water amplitude.*
*Glx (glutamate + glutamine)- tissue-corrected water-scaled.*
*Residual water amplitude in arbitrary units. CHR-P: Clinical High Risk for Psychosis.*

**Supplementary Table 7-** Seed-to-voxel analyses for the effect of anterior cingulate cortex (ACC) and hippocampus Glx levels on ACC and hippocampal functional connectivity.

| **Contrast** | Location | Peak T (40) | MNI coordinates (x,y,z) | Size (n voxels) | p-FWE | p-FDR |
|---|---|---|---|---|---|---|
| **ACC** | | | | | | |
| Effect Control ACC Glx | Nil significant | | | | | |
| Effect CHR ACC Glx | Middle temporal gyrus | -4.96 | -62 -16 -22 | 181 | 0.0083 | 0.0092 |
| Interaction group x ACC Glx | Nil significant | | | | | |
| **Hippocampus** | | | | | | |
| Effect Control ACC Glx | Nil significant | | | | | |
| Effect CHR ACC Glx | Left hippocampus, parahippocampal gyrus and temporal fusiform cortex | -4.96 | -30 -20 -24 | 177 | 0.0067 | 0.0078 |
| Interaction group x ACC Glx | Nil significant | | | | | |

Glx= glutamate + glutamine
Cluster-forming threshold of p<0.001, family-wise error (FWE) p-value and false detection rate (FDR for cluster size reported.

**Supplementary Table 8-** Pearson Correlation for anterior cingulate cortex (ACC) Glx levels and symptom scores split by group

| *Variable* | HC | | CHR-P | | FEP | |
|---|---|---|---|---|---|---|
| | **Pearson r** | **Pearson p-value** | **Pearson r** | **Pearson p-value** | **Pearson r** | **Pearson p-value** |
| Total IQ (WAIS-III) | -0.169 | 0.501 | 0.006 | 0.981 | -0.115 | 0.751 |
| NART | 0.143 | 0.571 | -0.407 | 0.105 | -0.504 | 0.167 |
| HAM-A | 0.115 | 0.650 | -0.072 | 0.777 | 0.108 | 0.766 |
| HAM-D | -0.428 | 0.077 | -0.156 | 0.537 | 0.322 | 0.364 |
| CAARMS Positive | 0.11 | 0.655 | -0.04 | 0.876 | - | - |
| CAARMS Negative | -0.156 | 0.628 | -0.336 | 0.173 | - | - |
| PANSS positive | - | - | - | - | 0.033 | 0.929 |
| PANSS negative | - | - | - | - | 0.318 | 0.371 |
| PANSS general | - | - | - | - | 0.496 | 0.145 |
| Trail Making A-B | -0.084 | 0.750 | 0.227 | 0.381 | 0.115 | 0.787 |
| WHODAS-II | 0.142 | 0.574 | -0.266 | 0.320 | 0.002 | 0.996 |
| SOFAS (Past Month) | -0.067 | 0.791 | 0.229 | 0.360 | -0.364 | 0.301 |

*Glx: tissue-corrected water-scaled glutamate + glutamine. HAM-A: Hamilton Anxiety Rating Scale. HAM-D: Hamilton Depression Rating Scale. CAARMS: Comprehensive Assessment of At-Risk Mental States. SOFAS: Social and Occupational Functioning Assessment Scale. WHODAS-II: World Health Organization Disability Assessment Schedule. WAIS-III: Wechsler Adult Intelligence Scale, 3rd Edition. NART: National Adult Reading Test.*

**Supplementary Table 9-** Pearson Correlation for left hippocampus Glx levels and symptom scores split by group

| Variable | HC | | CHR-P | | FEP | |
|---|---|---|---|---|---|---|
| | **Pearson r** | **Pearson p-value** | **Pearson r** | **Pearson p-value** | **Pearson r** | **Pearson p-value** |
| Total IQ (WAIS-III) | 0.316 | 0.201 | -0.003 | 0.989 | 0.135 | 0.709 |
| NART | 0.103 | 0.685 | 0.126 | 0.629 | 0.578 | 0.103 |
| HAM-A | -0.32 | 0.195 | 0.015 | 0.953 | 0.171 | 0.637 |
| HAM-D | 0.183 | 0.468 | 0.057 | 0.824 | -0.115 | 0.751 |
| CAARMS Positive | -0.05 | 0.840 | -0.17 | 0.500 | - | - |
| CAARMS Negative | 0.089 | 0.784 | 0.311 | 0.210 | - | - |
| PANSS Positive | - | - | - | - | -0.282 | 0.429 |
| PANSS Negative | - | - | - | - | -0.569 | 0.086 |
| PANSS General | - | - | - | - | -0.399 | 0.253 |
| Trail Making A-B | 0.302 | 0.238 | 0.016 | 0.951 | -0.685 | 0.061 |
| WHODAS-II | -0.151 | 0.549 | -0.015 | 0.956 | -0.35 | 0.442 |
| SOFAS (Past Month) | -0.368 | 0.133 | -0.248 | 0.321 | 0.538 | 0.109 |

*Glx: tissue-corrected water-scaled glutamate + glutamine. HAM-A: Hamilton Anxiety Rating Scale. HAM-D: Hamilton Depression Rating Scale. CAARMS: Comprehensive Assessment of At-Risk Mental States. SOFAS: Social and Occupational Functioning Assessment Scale. WHODAS-II: World Health Organization Disability Assessment Schedule. WAIS-III: Wechsler Adult Intelligence Scale, 3rd Edition. NART: National Adult Reading Test.*

**Supplementary Table 10-** Pearson Correlation for ACC ROI-to-ROI analyses beta values from Glx x group interaction and symptom scores for the clinical high risk for psychosis (CHR-P) group

| | **HAM-A** | **HAM-D** | **CAARMS Positive** | **CAARMS Negative** | **SOFAS** | **WHODAS-II** | **WAIS-III** | **NART** | **Trail making** |
|---|---|---|---|---|---|---|---|---|---|
| L NAc - L hippocampus | 0.027 (0.911) | -0.173 (0.466) | 0.189 (0.425) | -0.171 (0.471) | -0.031 (0.896) | 0.035 (0.889) | -0.117 (0.623) | -0.015 (0.953) | -0.221 (0.363) |
| L NAc – R amygdala | 0.213 (0.367) | 0.139 (0.559) | 0.037 (0.875) | -0.039 (0.872) | 0.117 (0.625) | 0.052 (0.836) | 0.155 (0.513) | 0.096 (0.695) | -0.194 (0.426) |
| L NAc – R hippocampus | 0.089 (0.709) | -0.050 (0.834) | 0.203 (0.391) | -0.240 (0.308) | -0.115 (0.629) | 0.280 (0.261) | -0.219 (0.354) | -0.264 (0.274) | -0.236 (0.330) |
| R NAc – L amygdala | 0.188 (0.427) | 0.362 (0.117) | -0.107 (0.655) | -0.238 (0.312) | -0.216 (0.361) | 0.221 (0.378) | -0.112 (0.639) | 0.152 (0.534) | -0.103 (0.675) |
| R NAc – L hippocampus | 0.156 (0.512) | 0.107 (0.654) | 0.056 (0.814) | -0.272 (0.245) | -0.110 (0.645) | 0.156 (0.537) | -0.120 (0.615) | 0.021 (0.933) | -0.348 (0.144) |
| R NAc – R amygdala | 0.296 (0.205) | 0.391 (0.0879) | 0.071 (0.765) | -0.139 (0.558) | -0.078 (0.743) | 0.043 (0.865) | 0.210 (0.374) | 0.200 (0.411) | -0.214 (0.378) |
| R NAc – R hippocampus | 0.186 (0.432) | 0.146 (0.538) | 0.122 (0.610) | -0.205 (0.385) | -0.187 (0.430) | 0.390 (0.110) | -0.060 (0.800) | 0.047 (0.848) | -0.247 (0.309) |

*Values are for pearson correlation r (p-value). Significant correlations are in* ***bold italic****. Glx: tissue-corrected water-scaled glutamate + glutamine. ROI: region-of-interest. NAc: nucleus accumbens. L: left, R: right. HAM-A: Hamilton Anxiety Rating Scale. HAM-D: Hamilton Depression Rating Scale. CAARMS: Comprehensive Assessment of At-Risk Mental States. SOFAS: Social and Occupational Functioning Assessment Scale. WHODAS-II: World Health Organization Disability Assessment Schedule. WAIS-III: Wechsler Adult Intelligence Scale, 3rd Edition. NART: National Adult Reading Test. Trail-making A-B score.*

**Supplementary Table 11-** Pearson Correlation for ACC ROI-to-ROI analyses beta values from Glx x group interaction and symptom scores for the health control (HC) group

| | **HAM-A** | **HAM-D** | **CAARMS Positive** | **CAARMS Negative** | **SOFAS** | **WHODAS-II** | **WAIS-III** | **NART** | **Trail making** |
|---|---|---|---|---|---|---|---|---|---|
| L NAc – L hippocampus | -0.010 (0.966) | -0.339 (0.155) | 0.067 (0.780) | 0.025 (0.935) | 0.063 (0.797) | 0.005 (0.984) | 0.318 (0.185) | 0.186 (0.445) | -0.045 (0.859) |
| L NAc – R amygdala | -0.138 (0.572) | -0.410 (0.081) | -0.086 (0.717) | -0.129 (0.674) | 0.066 (0.789) | -0.107 (0.674) | 0.074 (0.763) | 0.080 (0.744) | 0.001 (0.997) |
| L NAc – R hippocampus | 0.063 (0.797) | -0.306 (0.203) | 0.411 (0.0718) | -0.181 (0.553) | -0.140 (0.567) | -0.105 (0.680) | 0.037 (0.881) | -0.459 (0.0479) | 0.254 (0.309) |
| R NAc – L amygdala | -0.284 (0.239) | ***-0.508 (0.0265)*** | 0.173 (0.466) | 0.249 (0.412) | -0.351 (0.140) | -0.044 (0.862) | -0.002 (0.993) | 0.054 (0.825) | 0.019 (0.941) |
| R NAc – L hippocampus | -0.051 (0.836) | -0.057 (0.817) | 0.044 (0.856) | 0.241 (0.428) | -0.442 (0.058) | 0.130 (0.608) | 0.057 (0.816) | 0.368 (0.121) | 0.040 (0.873) |
| R NAc – R amygdala | -0.388 (0.101) | ***-0.520 (0.0226)*** | -0.189 (0.425) | -0.260 (0.392) | -0.186 (0.445) | -0.219 (0.382) | -0.111 (0.651) | 0.053 (0.828) | 0.175 (0.488) |
| R NAc – R hippocampus | 0.038 (0.877) | -0.004 (0.986) | 0.163 (0.493) | 0.309 (0.304) | -0.546 (0.0156) | 0.117 (0.643) | 0.030 (0.902) | -0.204 (0.402) | 0.260 (0.297) |

*Values are for pearson correlation r (p-value). Significant correlations are in* ***bold italic****. Glx: tissue-corrected water-scaled glutamate + glutamine. ROI: region-of-interest. NAc: nucleus accumbens. L: left, R: right. HAM-A: Hamilton Anxiety Rating Scale. HAM-D: Hamilton Depression Rating Scale. CAARMS: Comprehensive Assessment of At-Risk Mental States. SOFAS: Social and Occupational Functioning Assessment Scale. WHODAS-II: World Health Organization Disability Assessment Schedule. WAIS-III: Wechsler Adult Intelligence Scale, 3rd Edition. NART: National Adult Reading Test. Trail-making A-B score.*

### 1 Supplementary Figures

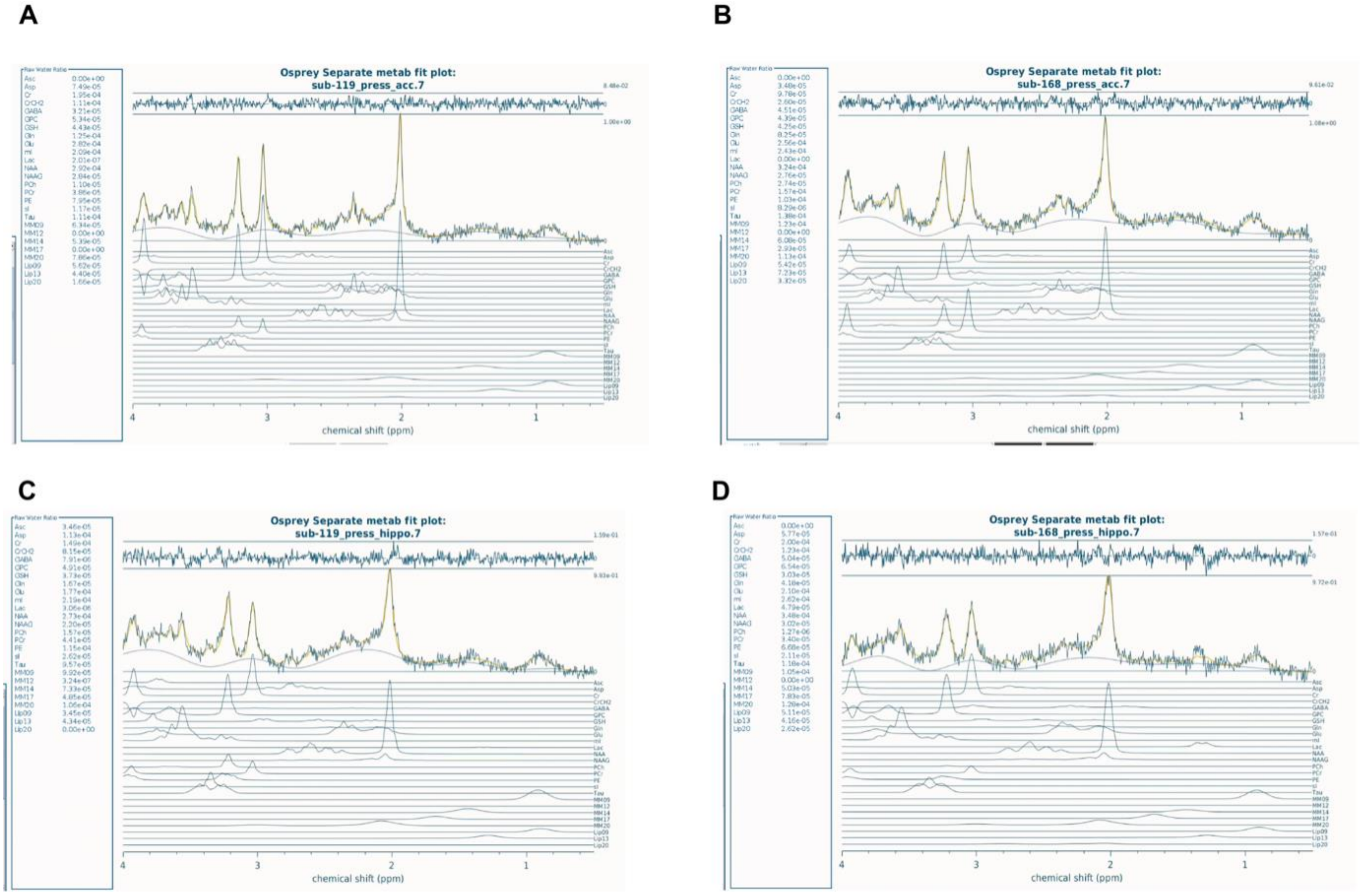

**Supplementary Figure 1- Sample spectra from Osprey for anterior cingulate cortex (ACC) and left hippocampus 1H-MRS PRESS for two sample participants.** (A-B) Sample spectra for ACC MRS for two participants. (C-D) Sample spectra for left hippocampus MRS for two participants.

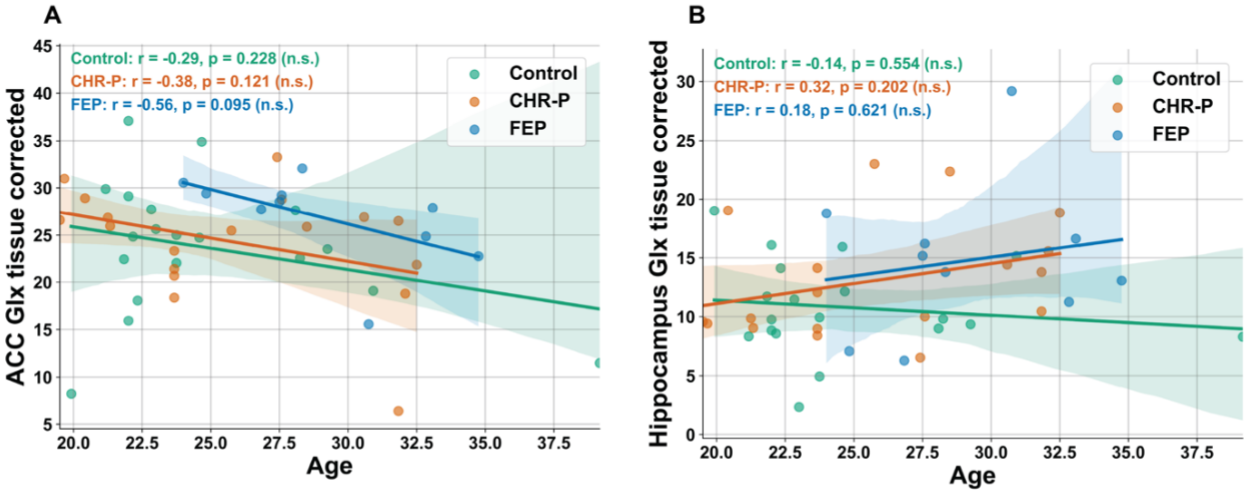

**Supplementary Figure 2- Association between anterior cingulate cortex (ACC) and left hippocampus $^{1}$H-MRS Glx levels and age compared by group: healthy controls (HC), clinical high risk for psychosis (CHR-P) and first episode psychosis (FEP).** Scatter plot with regression lines with Pearson correlation (2-tailed) values and p-values for ACC Glx and age (A) and for left hippocampus Glx and age (B). Shaded areas around regression lines represent 95% confidence intervals. n.s= nonsignificant.

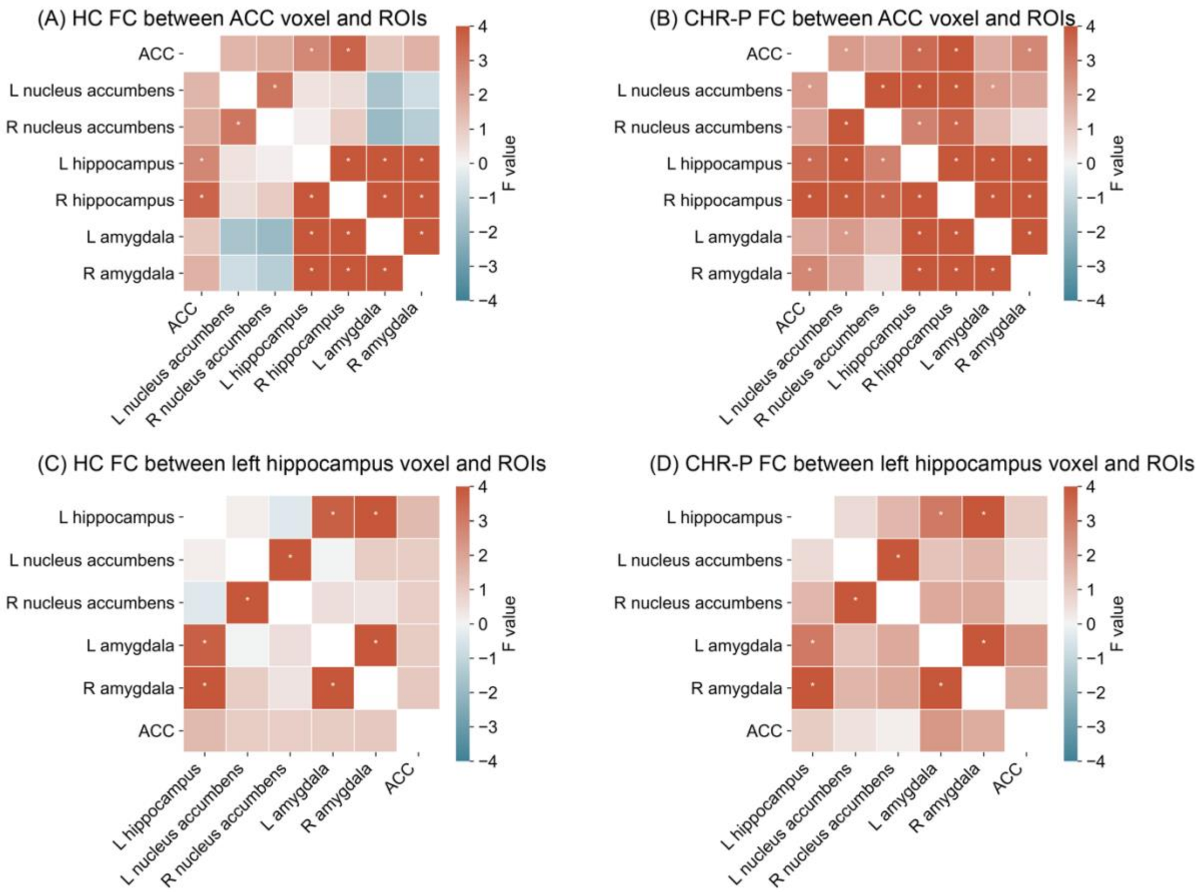

**Supplementary Figure 3- ROI-to-ROI functional connectivity networks for anterior cingulate cortex (ACC) and left hippocampus averaged across each group** Average ACC ROI-to-ROI functional connectivity for the control group (A) and clinical high risk for psychosis (CHR-P) group (B). Average hippocampal ROI-to-ROI functional connectivity for the control group (C) and clinical high risk for psychosis (CHR-P) group (D). Red/ yellow colours indicate positive T-values and blue indicated negative T-values. Asterix indicates a significant connection after FDR correction.

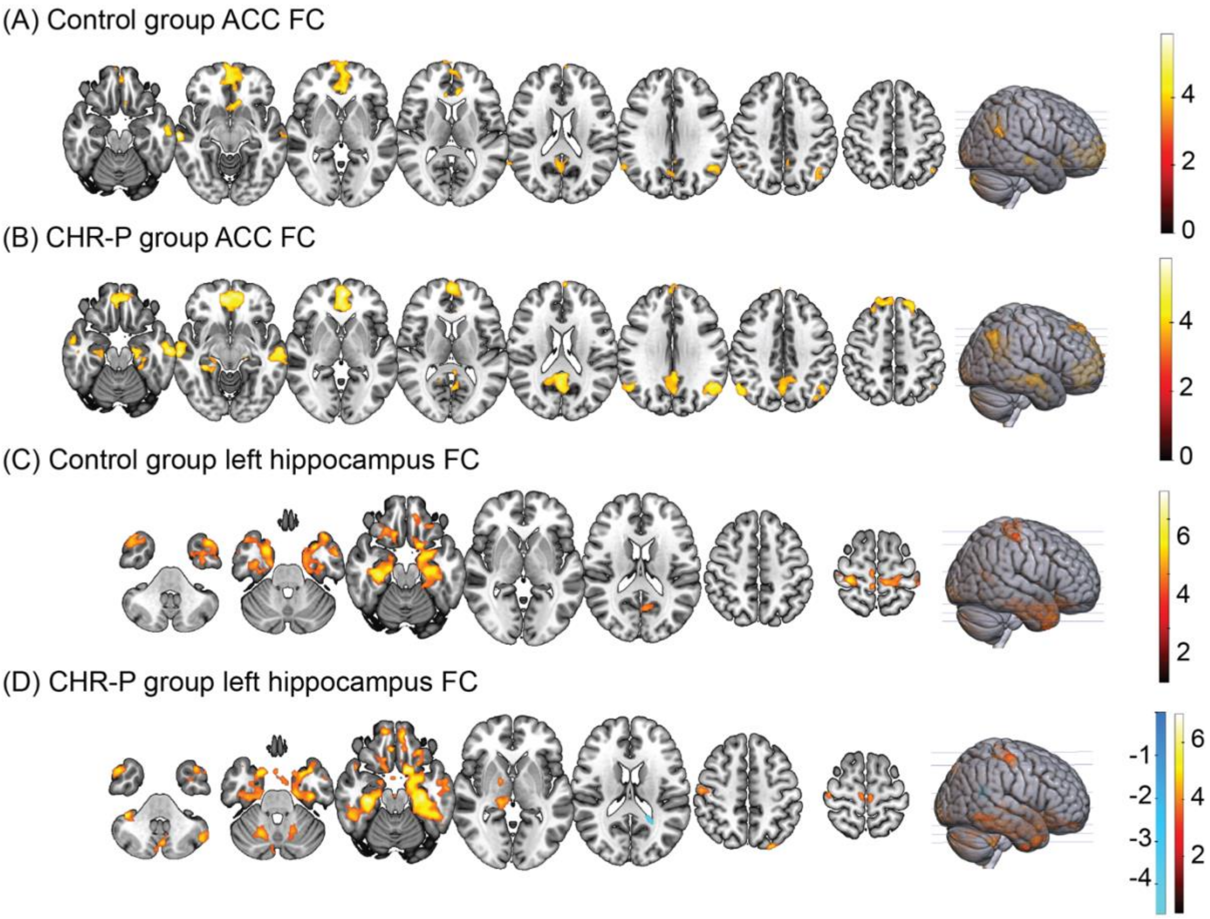

**Supplementary Figure 4- Seed-to-voxel functional connectivity networks for anterior cingulate cortex (ACC) and left hippocampus averaged across each group** Average ACC seed-to-voxel functional connectivity for the control group (A) and clinical high risk for psychosis (CHR-P) group (B). Average hippocampal seed-to-voxel functional connectivity for the control group (C) and clinical high risk for psychosis (CHR-P) group (D). Red/ yellow colours indicate positive T-values and blue indicated negative T-values.
